# Monitoring global trends in Covid-19 vaccination intention and confidence: a social media-based deep learning study

**DOI:** 10.1101/2021.04.17.21255642

**Authors:** Xinyu Zhou, Alex de Figueiredo, Qin Xu, Leesa Lin, Per E Kummervold, Heidi Larson, Mark Jit, Zhiyuan Hou

**Affiliations:** School of Public Health, Global Health Institute, NHC Key Laboratory of Health Technology Assessment, Fudan University, Shanghai, China; Department of Infectious Disease Epidemiology, London School of Hygiene & Tropical Medicine, UK; FISABIO-Public Health, Vaccine Research Department, Valencia, Spain

**Keywords:** Covid-19, vaccine, acceptance, intention, confidence, social media, machine learning

## Abstract

**Background:** This study developed deep learning models to monitor global intention and confidence of Covid-19 vaccination in real time.

**Methods:** We collected 6.73 million English tweets regarding Covid-19 vaccination globally from January 2020 to February 2021. Fine-tuned Transformer-based deep learning models were used to classify tweets in real time as they relate to Covid-19 vaccination intention and confidence. Temporal and spatial trends were performed to map the global prevalence of Covid-19 vaccination intention and confidence, and public engagement on social media was analyzed.

**Findings:** Globally, the proportion of tweets indicating intent to accept Covid-19 vaccination declined from 64.49% on March to 39.54% on September 2020, and then began to recover, reaching 52.56% in early 2021. This recovery in vaccine acceptance was largely driven by the US and European region, whereas other regions experienced the declining trends in 2020. Intent to accept and confidence of Covid-19 vaccination were relatively high in South-East Asia, Eastern Mediterranean, and Western Pacific regions, but low in American, European, and African regions. 12.71% tweets expressed misinformation or rumors in South Korea, 14.04% expressed distrust in government in the US, and 16.16% expressed Covid-19 vaccine being unsafe in Greece, ranking first globally. Negative tweets, especially misinformation or rumors, were more engaged by twitters with fewer followers than positive tweets.

**Interpretation:** This global real-time surveillance study highlights the importance of deep learning based social media monitoring to detect emerging trends of Covid-19 vaccination intention and confidence to inform timely interventions.

**Funding:** National Natural Science Foundation of China.

**Research in context:** *Evidence before this study:* With COVID-19 vaccine rollout, each country should investigate its vaccination intention in local contexts to ensure massive vaccination. We searched PubMed for all articles/preprints until April 9, 2021 with the keywords “(“Covid-19 vaccines”[Mesh] OR Covid-19 vaccin*[TI]) AND (confidence[TI] OR hesitancy[TI] OR acceptance[TI] OR intention[TI])”. We identified more than 100 studies, most of which are country-level cross-sectional surveys, and the largest global survey of Covid-19 vaccine acceptance only covered 32 countries to date. However, how Covid-19 vaccination intention changes over time remain unknown, and many countries are not covered in previous surveys yet. A few studies assessed public sentiments towards Covid-19 vaccination using social media data, but only targeting limited geographical areas. There is a lack of real-time surveillance, and no study to date has globally monitored Covid-19 vaccination intention in real time.

*Added value of this study:* To our knowledge, this is the largest global monitoring study of Covid-19 vaccination intention and confidence with social media data in over 100 countries from the beginning of the pandemic to February 2021. This study developed deep learning models by fine-tuning a Bidirectional Encoder Representation from Transformer (BERT)-based model with 8000 manually-classified tweets, which can be used to monitor Covid-19 vaccination beliefs using social media data in real time. It achieves temporal and spatial analyses of the evolving beliefs to Covid-19 vaccines across the world, and also an insight for many countries not yet covered in previous surveys. This study highlights that the intention to accept Covid-19 vaccination have experienced a declining trend since the beginning of the pandemic in all world regions, with some regions recovering recently, though not to their original levels. This recovery was largely driven by the US and European region (EUR), whereas other regions experienced the declining trends in 2020. Intention to accept and confidence of Covid-19 vaccination were relatively high in South-East Asia region (SEAR), Eastern Mediterranean region (EMR), and Western Pacific region (WPR), but low in American region (AMR), EUR, and African region (AFR). Many AFR countries worried more about vaccine effectiveness, while EUR, AMR, and WPR concerned more about vaccine safety (the most concerns with 16.16% in Greece). Online misinformation or rumors were widespread in AMR, EUR, and South Korea (12.71%, ranks first globally), and distrust in government was more prevalent in AMR (14.04% in the US, ranks first globally). Our findings can be used as a reference point for survey data on a single country in the future, and inform timely and specific interventions for each country to address Covid-19 vaccine hesitancy.

*Implications of all the available evidence:* This global real-time surveillance study highlights the importance of deep learning based social media monitoring as a quick and effective method for detecting emerging trends of Covid-19 vaccination intention and confidence to inform timely interventions, especially in settings with limited sources and urgent timelines. Future research should build multilingual deep learning models and monitor Covid-19 vaccination intention and confidence in real time with data from multiple social media platforms.

## Introduction

Large-scale uptake of Covid-19 vaccines will be crucial for pandemic control, yet reluctance to vaccinate in a number of global settings will pose challenges for herd immunity targets.^1^ A 32-country survey on potential acceptance of Covid-19 vaccines found high heterogeneity across countries, ranging from more than 90% (in Vietnam, India, and China) to around 40% (in France, Serbia, and Croatia) in late 2020.^1^ With Covid-19 vaccine rollout, each country should investigate Covid-19 vaccination acceptance in local contexts to ensure massive vaccination. However, many countries with low acceptance have not been identified ^2,3^, and there is a paucity of research on the evolution in Covid-19 vaccination acceptance over time^4^.

Previous studies on Covid-19 vaccine acceptance have predominantly relied on cross-sectional questionnaire-based surveys^1-3^. Recent years, however, social media has been increasingly recognized as a platform to understand and respond to public attitudes and behaviours, especially during public health emergencies^5,6^. Compared to traditional surveys, social media platforms could provide real-time data to monitor the public’s attitudes to vaccines in large scales^7,8^. A growing body of literature has used social media for vaccination attitude research^6^, and a few studies assessed sentiments towards Covid-19 vaccination using social media data, but only targeting limited geographical areas.^9-12^ There is a lack of real-time surveillance, and no study to date has globally monitored Covid-19 vaccine acceptance with social media data.^1,12^

The introduction of transformer-based deep learning model has improved performance in various natural language processing tasks^13,14^, thus making it possible to analyze massive textual data on social media with high accuracy in real time. In this study, we analyzed all publicly available English tweets regarding Covid-19 vaccination from the beginning of Covid-19 outbreak to February 2021, using deep learning models. We aimed to assess the global temporal and spatial trends of Covid-19 vaccination intention and confidence in real time, and compare the propagation of various contents about Covid-19 vaccination on social media. This global real-time surveillance study can detect emerging trends of vaccination intention and confidence to inform potential countermeasures against Covid-19 vaccine hesitancy.

## Methods

### Data collection

We collected around 6.73 million English-language tweets regarding Covid-19 vaccination across the globe from January 1, 2020 to February 28, 2021 through Twitter advanced search (https://twitter.com/search-advanced) using TweetScraper (https://github.com/jonbakerfish/TweetScraper) on Google Cloud (Ubuntu 20.04). To assess intention and confidence of Covid-19 vaccination, we only included tweets from private accounts, excluding tweets from news media, organizational accounts, or verified users; duplicate tweets and tweets with identical text but different tweet identifications were also removed, leading to a preprocessed dataset of ∼5.04 million tweets (appendix 1 pp 1). This study was exempt from ethical review due to use of retrospective, publicly available data. An overview of study design is shown in Figure 1, and some details in appendix 1.

**Figure 1.**
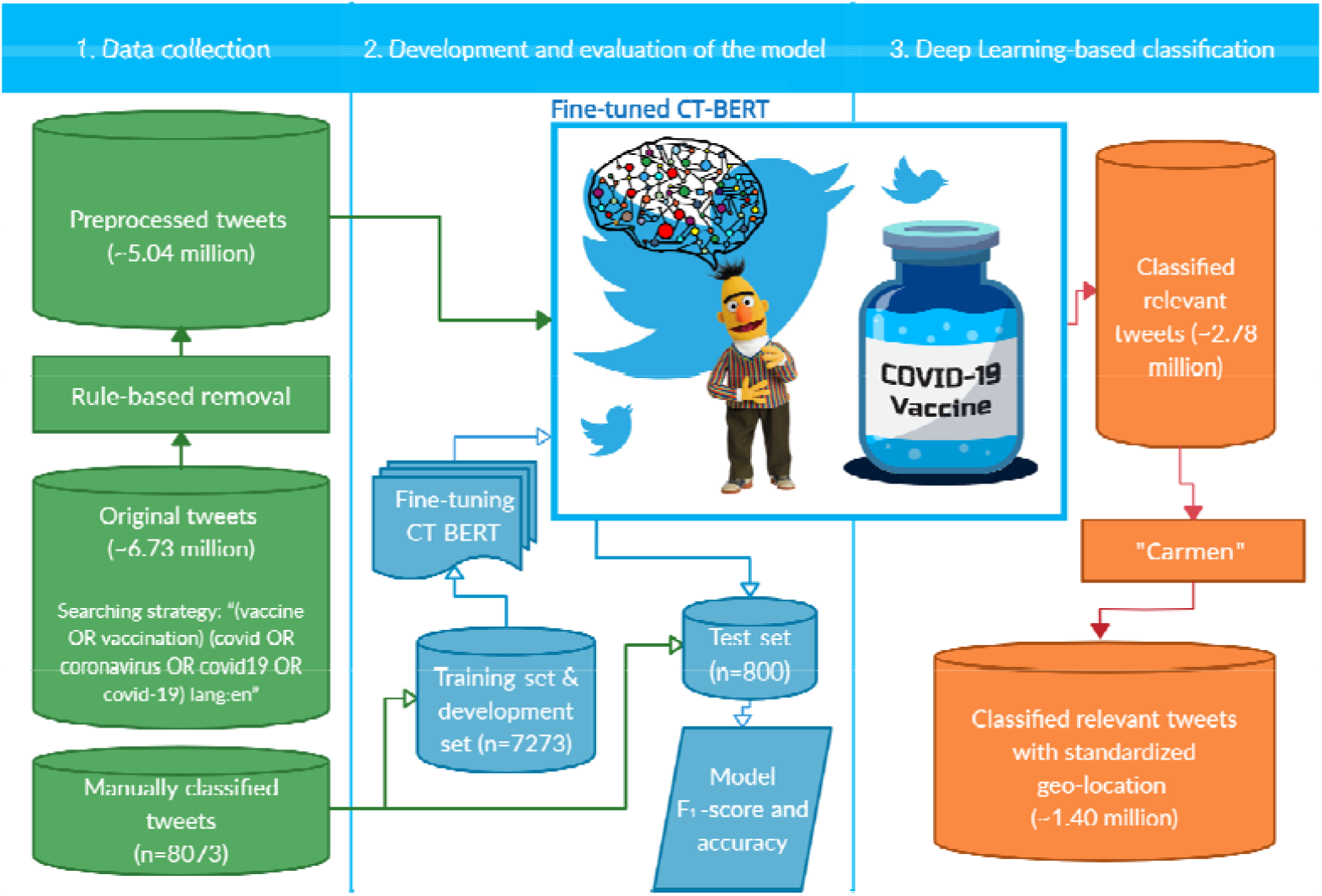
Flowchart of the methodology.

### Development of deep learning model

Bidirectional Encoder Representation from Transformer (BERT), proposed by Google in 2018, is a 12/24-layer deep neural network with 110/340 million parameters that outperformed all models before it on the General Language Understanding Evaluation benchmark, with 7.0% absolute improvement over the prior state of the art model.^13,14^ By pre-training multi-layer bidirectional Transformer encoder with BooksCorpus (800 million words) and English Wikipedia (2,500 million words) to better “understand” a language, BERT can be applied to classify textual data by fine-tuning (aka. adjusting parameters) with a limited manually-classified downstream training set.^13,14^ COVID-Twitter-BERT (CT-BERT)^13,16^, a model based on the 24-layers BERT and further pretrained with 160 million Covid-19 related tweets, is reported to increase BERT’s marginal performance by 10%-30% in tweet-like or Covid-19 related data. ^16^ In this study, deep learning models were developed by fine-tuning CT-BERT with manually-classified tweets regarding Covid-19 vaccination using Google Colab.

First, around 8000 tweets were manually classified to predefined categories according to the framework of vaccine hesitancy proposed by the World Health Organization (Table 1)^17,18,19^. Each tweet was classified by two independent annotators, and a third annotator resolved disagreements. Second, the initially manually classified tweets were used to train, validate, and test deep learning models. We randomly selected 80% of the 8000 classified tweets as the training set, 10% as the validation set, and 10% as the test set. The training set and validation set were used to finetune and validate the deep neural networks. Models with unsatisfactory performance in the validation set were excluded, and models included in this study was tested on the remaining 10% classified tweets using two widely-used indicators - accuracy and f_1_-score (the harmonic mean of accuracy and recall, widely used in machine learning; see appendix 1 pp 2). In the test set, the model for identifying whether tweets were relevant or not to Covid-19 vaccination intention and confidence reached an accuracy of 89% (f_1_-score=0.86), and the model for further classifying relevant tweets to predefined categories reached an accuracy of 82% (f_1_-score=0.81) (Table 1).

**Table 1.**
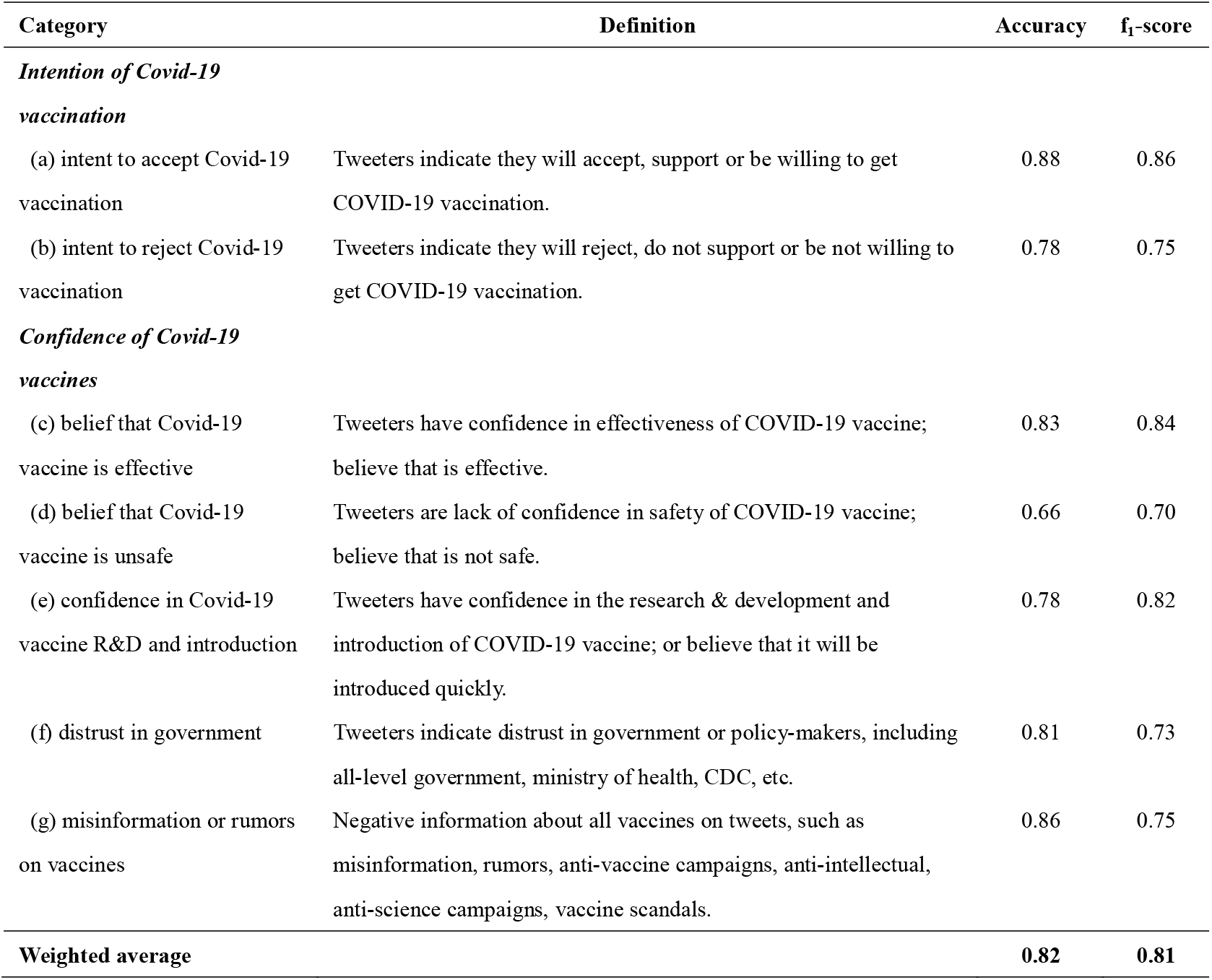
Classification framework and the performance of deep learning models.

### Deep learning-based classification of tweets

We first identified the relevant tweets dataset (n=2,781,591) from the preprocessed dataset, and further classified the relevant tweets with deep learning models. Using the fine-tuned deep learning models, relevant tweets were classified to the predefined categories in Table 1. First, each tweet would be classified as one of the following categories on Covid-19 vaccination intention: (a) intent to accept Covid-19 vaccination, or (b) intent to reject Covid-19 vaccination, or others (neutral or cannot judge). Second, tweets would be classified as some of the following categories on Covid-19 vaccine confidence: (c) belief that Covid-19 vaccine is effective, (d) belief that Covid-19 vaccine is unsafe, (e) confidence in Covid-19 vaccine R&D and introduction, (f) distrust in government, or (g) misinformation or rumors on vaccines; each tweet was classified into one category, multiple categories, or no category regarding vaccine confidence. Geo-location of tweets were collected using Carmen (https://github.com/mdredze/carmen-python/tree/master/carmen),^15^ which provided standardized locations for half of the tweets (n=1,400,492) by simultaneously utilizing GPS data and Twitter user’s profile location.

### Statistical analysis

We defined the prevalence of each category regarding Covid-19 vaccination intention and confidence as the proportion of tweets that were classified to that category among all relevant tweets. We conducted the temporal and spatial analyses of Covid-19 vaccination intention and confidence across the world in real time. Temporal trend for each category was estimated by its prevalence each day, month, and three-months (season). Spatial variations were estimated across world regions or countries for tweets with standardized geo-locations.

We conducted univariate linear regressions between (a) intent to accept Covid-19 vaccination and the remaining six categories (b-g) using country-level data. We also validated our tweet data with the previous global survey data on Covid-19 vaccine acceptance using univariate linear regression.^1^ We further compared public engagement on Twitter across different categories by measuring the mean and standard deviation of followers, retweets, favorites, replies, and quote retweets for all tweets within each category. Data was analyzed with Python.

## Results

Globally, around 2.78 million English tweets relevant to Covid-19 vaccination intention and confidence were posted by individuals between January 2020 and February 2021, of which around 1.40 million tweets had standardized geo-location data. Among geo-located tweets, the United States accounted for the largest proportion (788,043, 56.27 %), followed by the United Kingdom (219,522, 19.14 %), Canada (66,438, 5.38 %), India (58,470, 4.52 %), and Australia (31,166, 2.64%). Vaccine-related discussions increased with newly confirmed Covid-19 cases (Figure 2). There were also abrupt increases when major developments regarding Covid-19 vaccine were announced, e.g. a spike on November 9 in tweets when Pfizer/BioNTech announced the results of phase III vaccine clinical trial, and a spike on November 16 when Moderna announced the preliminary data from their phase III trial.

**Figure 2.**
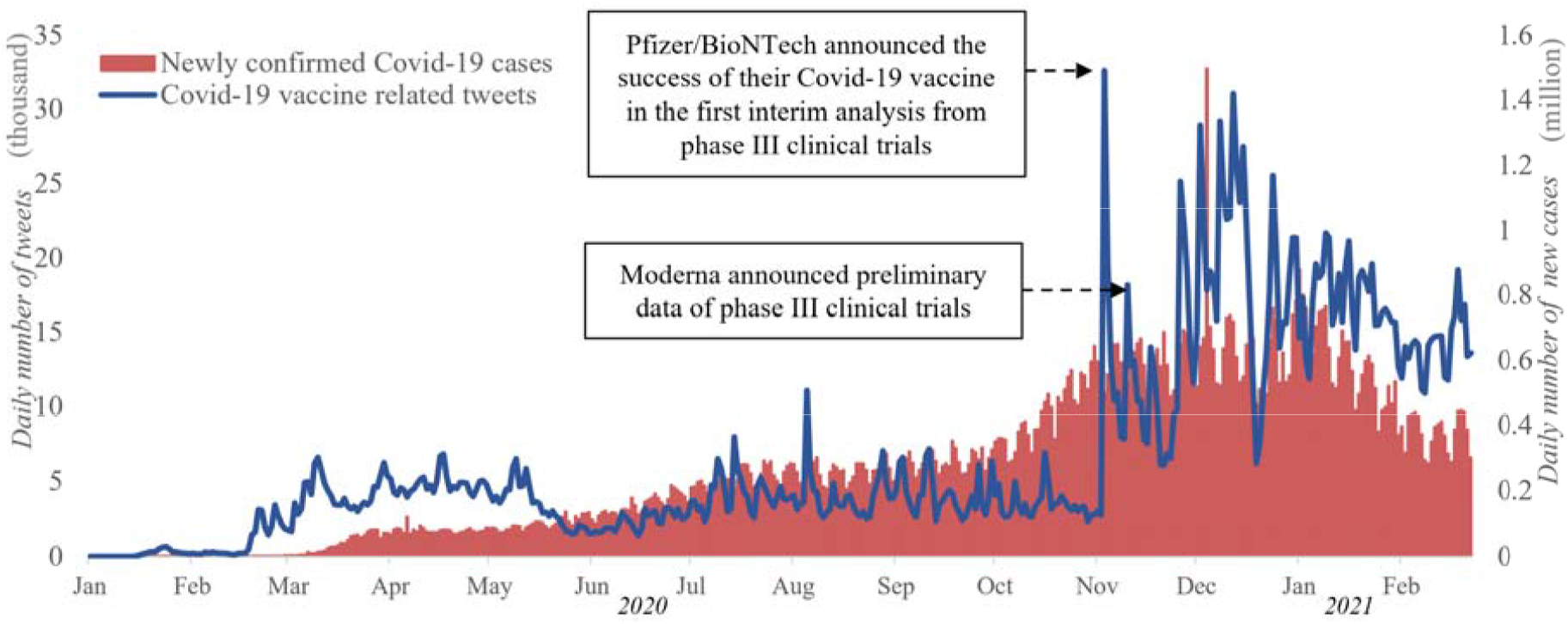
Daily number of Covid-19 vaccine related tweets and new Covid-19 cases between January 2020 and February 2021.

Figure 3 presents temporal trends for Covid-19 vaccination intention and confidence globally and by WHO region, and appendix figure 1 presents temporal trends in selected countries. Globally, the proportion of tweets that indicated intent to accept Covid-19 vaccination declined from a daily average of 64.49% on March to 39.54% on September 2020, and then began to recover, reaching a daily average of 52.56% between January 1 to February 28, 2021 (Figure 3a).

**Figure 3.**
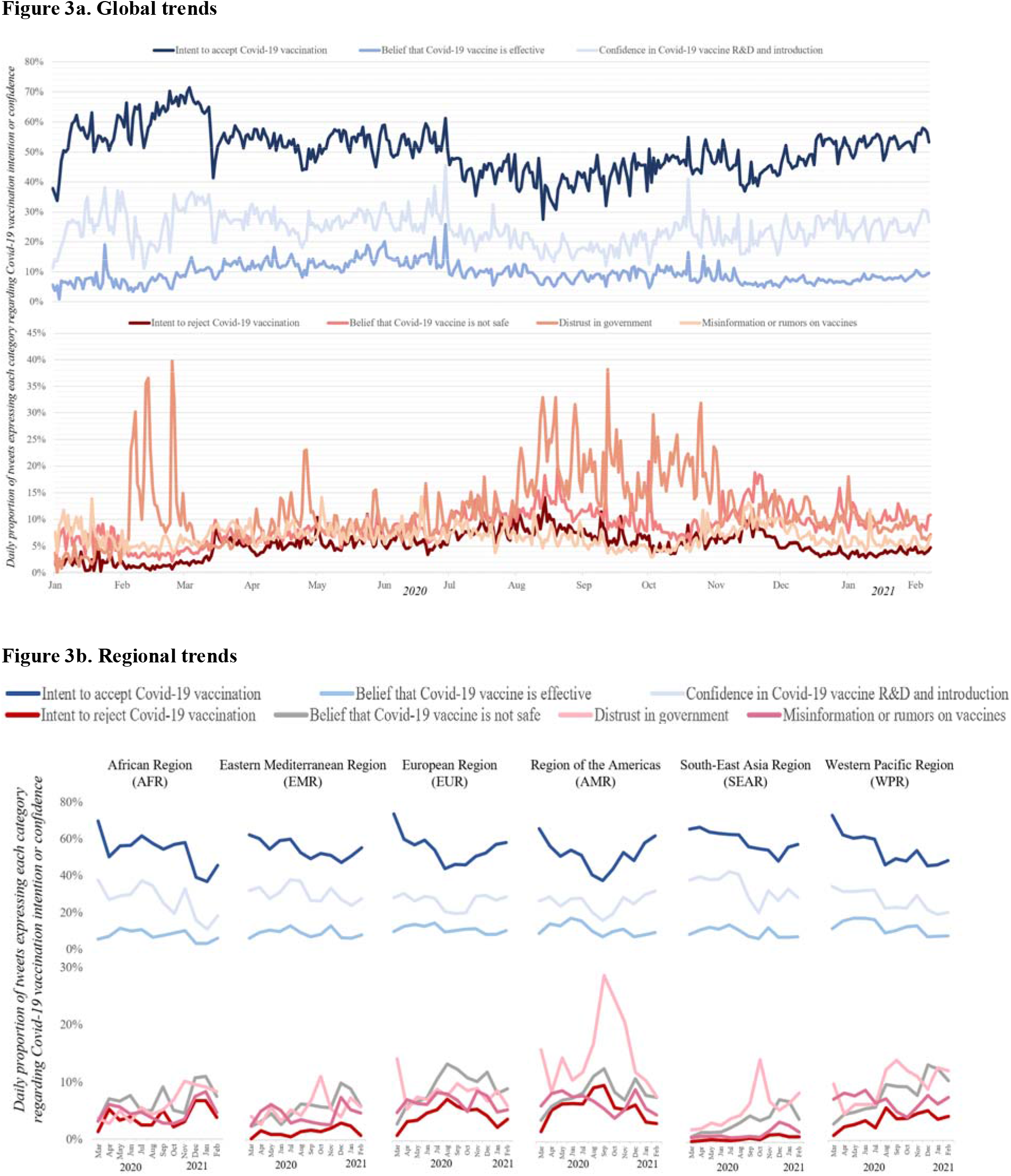
Temporal trends in Covid-19 vaccination intention and confidence between January 2020 and February 2021.

The proportion of intent to accept Covid-19 vaccination varied across WHO’s world regions, but fell consistently in the African region (AFR), Eastern Mediterranean region (EMR), South-East Asia region (SEAR), and Western Pacific region (WPR) during the whole year of 2020 (Figure 3b). However, in the European region (EUR) and American region (AMR), after a decrease in the proportion of tweets indicating intent to accept Covid-19 vaccination in the first half of 2020, the proportion grew from September 2020. Trends in perceptions towards the effectiveness of Covid-19 vaccines and in confidence around vaccine R&D and introduction largely mirrored those for vaccine acceptance with either persistent declines (AFR, EMR, SEAR, WPR) or an initial decline followed by a recovery in the latter half of 2020 (EUR, AMR) (Figure 3b). In contrast, the proportions of tweets indicating intent to reject Covid-19 vaccination or belief that Covid-19 vaccine is unsafe have risen fairly consistently in AFR and WPR, but some recent reversal in negative perceptions is evident in the other WHO regions. The proportion of tweets indicating distrust in government experienced an abrupt increase between August to October 2020 when Covid-19 resurged in many countries, especially in AMR. The proportion of tweets containing misinformation or rumors about vaccines fluctuated slightly between January 2020 to February 2021.

Figure 4 and appendix figure 2 reveal global comparisons of Covid-19 vaccination intention and confidence. On average, tweets located in SEAR, EMR and WPR tended to accept Covid-19 vaccine and expressed more confidence in Covid-19 vaccine R&D and introduction than tweets in AMR, EUR and AFR (appendix figure 2). SEAR and EMR dominate the top-20 list of countries with acceptance of Covid-19 vaccines, including five countries from SEAR (Bangladesh, 76.00%, ranks first globally; India, 72.29%; Maldives, 68.58%; Nepal, 68.38%; Sri Lanka, 64.39%) and seven countries from EMR (Oman, 70.37%; Pakistan, 69.43%; United Arab Emirates, 68.06%; Qatar, 67.90%; Iraq, 66.39%; Afghanistan, 65.90%; Saudi Arabia, 64.20%) (appendix 2). However, of the 20 countries with the lowest acceptance of Covid-19 vaccines, seven are from EUR (Latvia, 35.94%; Monaco, 36.17%; Serbia, 39.77%; Greece, 43.78%; Iceland, 44.08%; Russia, 44.34%; Slovenia, 48.00%), five are from AMR (Bahamas, 36.69%; Barbados, 40.89%; Chile, 47.26%; Jamaica, 47.44%; Costa Rica, 47.47%), and four are from AFR (Namibia, 43.35%; South Africa, 43.53%; Malawi, 46.54%; Cameroon, 47.68%) (appendix 2). The proportion of tweets indicating a belief that Covid-19 vaccine is unsafe was higher in EUR, AMR, and WPR than other regions (appendix figure 2), with the highest proportion of tweets questioning the safety of Covid-19 vaccines in Greece (16.16%, ranks first globally). Seven of eight countries with the lowest proportion of tweets indicating Covid-19 vaccine being effective were from AFR (Ghana, Nigeria, Kenya, South Africa, Namibia, Zimbabwe, and Uganda) (appendix 2). Distrust in government was more prevalent in tweets from AMR (appendix figure 2), with the most prevalent distrust in the United States (14.04%, ranks first globally). Online misinformation or rumors were widespread in AMR, EUR, and South Korea. Note that in South Korea, the proportion of tweets indicating intent to accept Covid-19 vaccination was low (42.27%), and proportions of tweets indicating intent to reject Covid-19 vaccination (7.73%) and misinformation (12.71%, ranks first globally) were high. The proportions of tweets expressing each category in Covid-19 vaccination intention and confidence by country are presented in appendix figure 3 and appendix 2.

**Figure 4.**
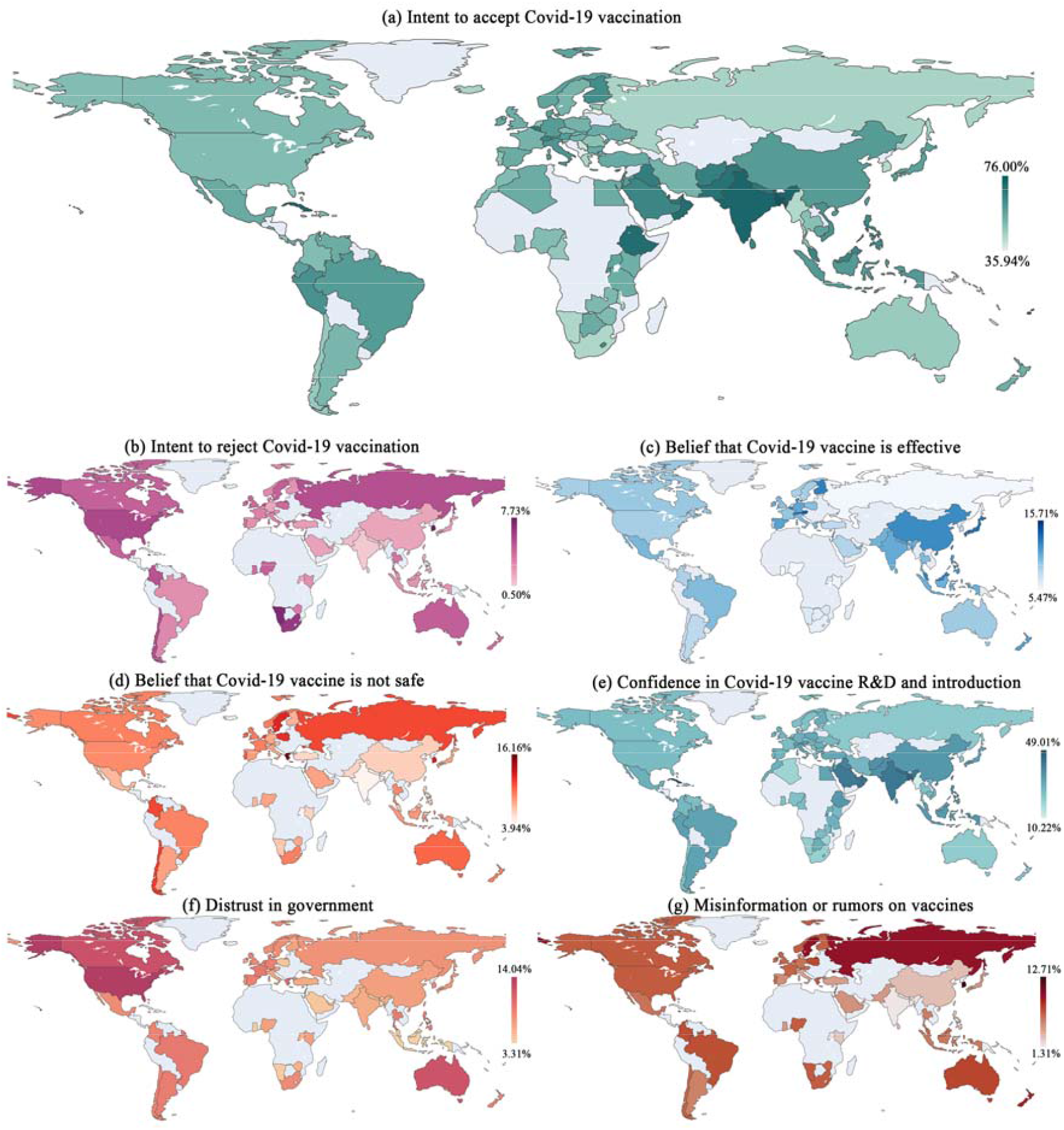
Global comparison of Covid-19 vaccination intention and confidence between January 2020 and February 2021. At the country level, the graphs below illustrate the proportion of tweets which contain the following categories regarding Covid-19 vaccination intention or confidence. Note: • Only countries with >100 relevant tweets are presented in: (a) intent to accept Covid-19 vaccination (e) confidence in Covid-19 vaccine R&D and introduction. • Only countries with >500 relevant tweets are presented in: (b) intent to reject Covid-19 vaccination, (c) belief that Covid-19 vaccine is effective, (d) belief that Covid-19 vaccine is unsafe, (f) distrust in government, and (g) misinformation or rumors on vaccines. • Countries without adequate data are in grey.

We also mapped the temporal trends of Covid-19 vaccination intention and confidence in each country in every three-months (seasons) to ensure enough data (Figure 5). This figure highlighted a decline in the proportion of tweets indicating intent to accept Covid-19 vaccination averaged across all countries (65.18% during March to May 2020 (spring) →57.54% during June to August 2020 (summer) →55.55% during September to November 2020 (autumn) →52.78% during December 2020 to February 2021 (winter)), and a small raise in the proportion of tweets indicating intent to reject Covid-19 vaccination (2.57%→3.03%→3.20%→3.37%). Out of 55 countries, the proportion of tweets indicating intent to accept Covid-19 vaccination dropped from spring to summer in 45 countries, from summer to autumn in 37 countries, and from autumn to winter in 35 countries. Countries such as South Korea, Turkey, Netherlands, and Greece experienced a large drop in intent to accept Covid-19 vaccination between spring and summer 2020, but recovered in autumn 2020. The proportion of intent to accept Covid-19 vaccination recovered in winter in the US, UK, Ireland, Denmark, Austria, Russia, and the United Arab Emirates, although not recovering to their original levels. In contrast, countries in AFR such as Uganda, Nigeria, Kenya and Zimbabwe bucked the global trend - the proportion of intent to accept Covid-19 vaccination rose from spring to summer, but experienced a large drop in winter, 2020. Monthly changes of proportions of intent to accept Covid-19 vaccination in 22 selected countries between November 2020 and February 2021 are presented in appendix figure 4, showing that after a drop from November 2020 to December 2020, Covid-19 vaccine acceptance was recovering during early 2021.

**Figure 5.**
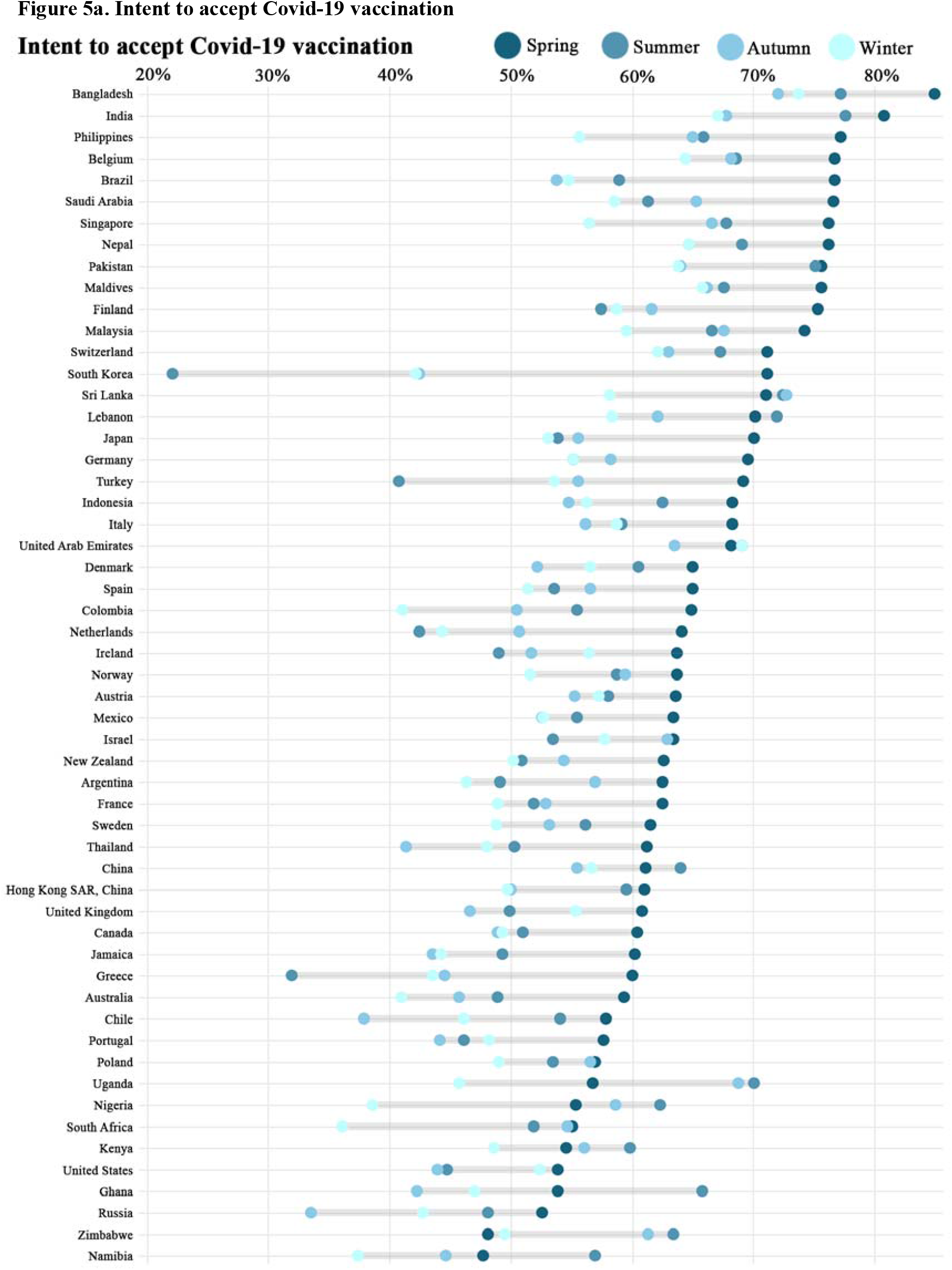
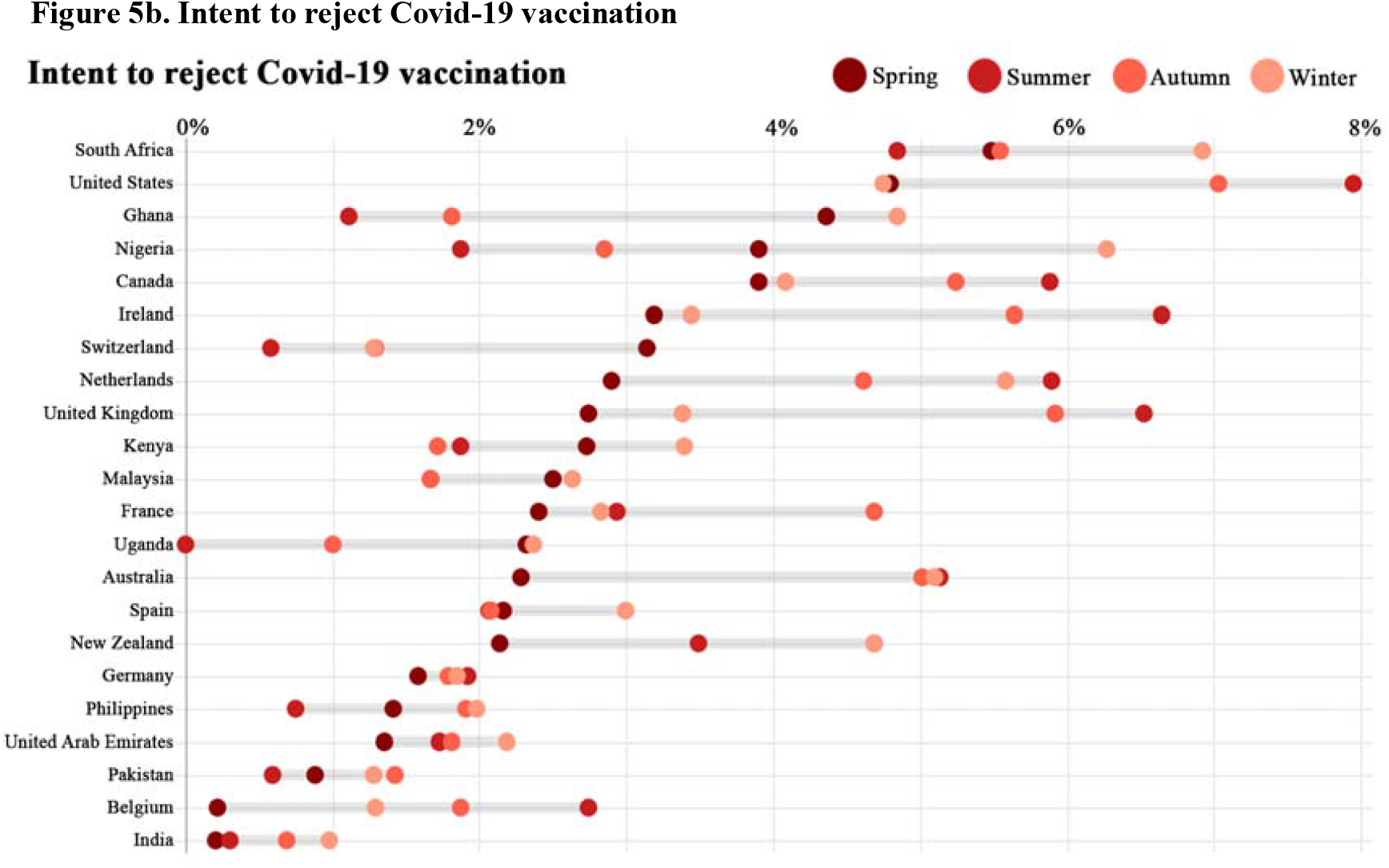
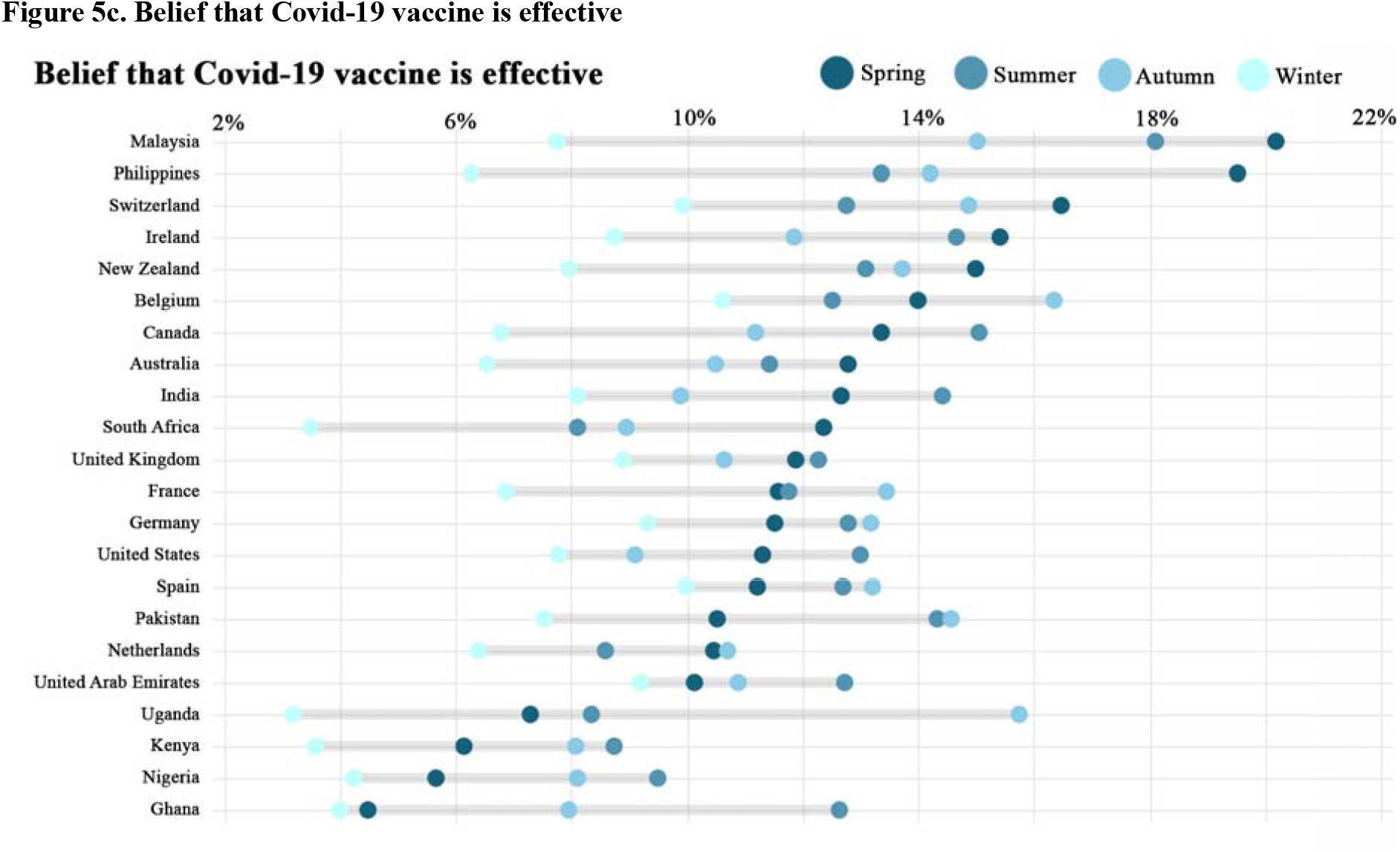
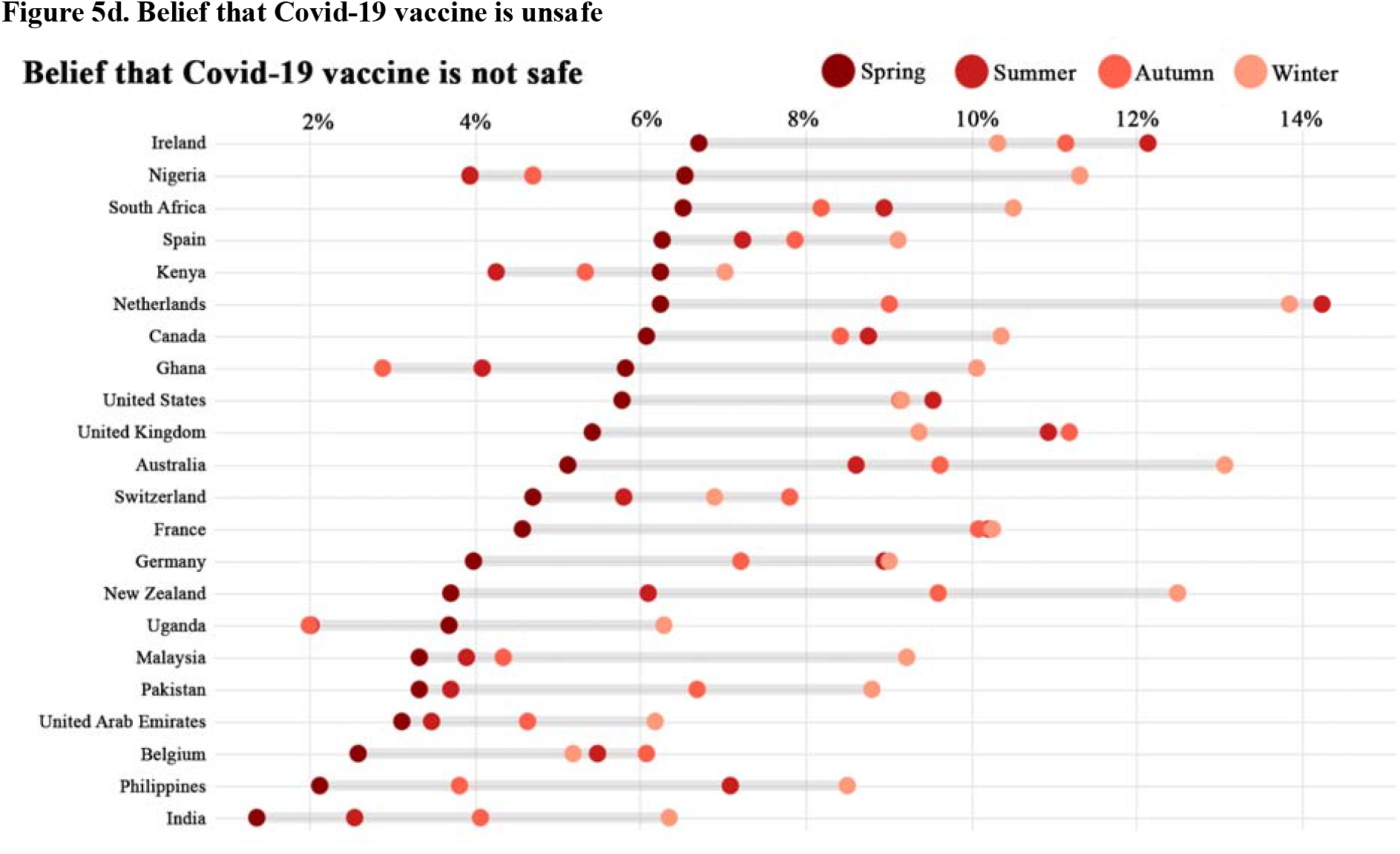
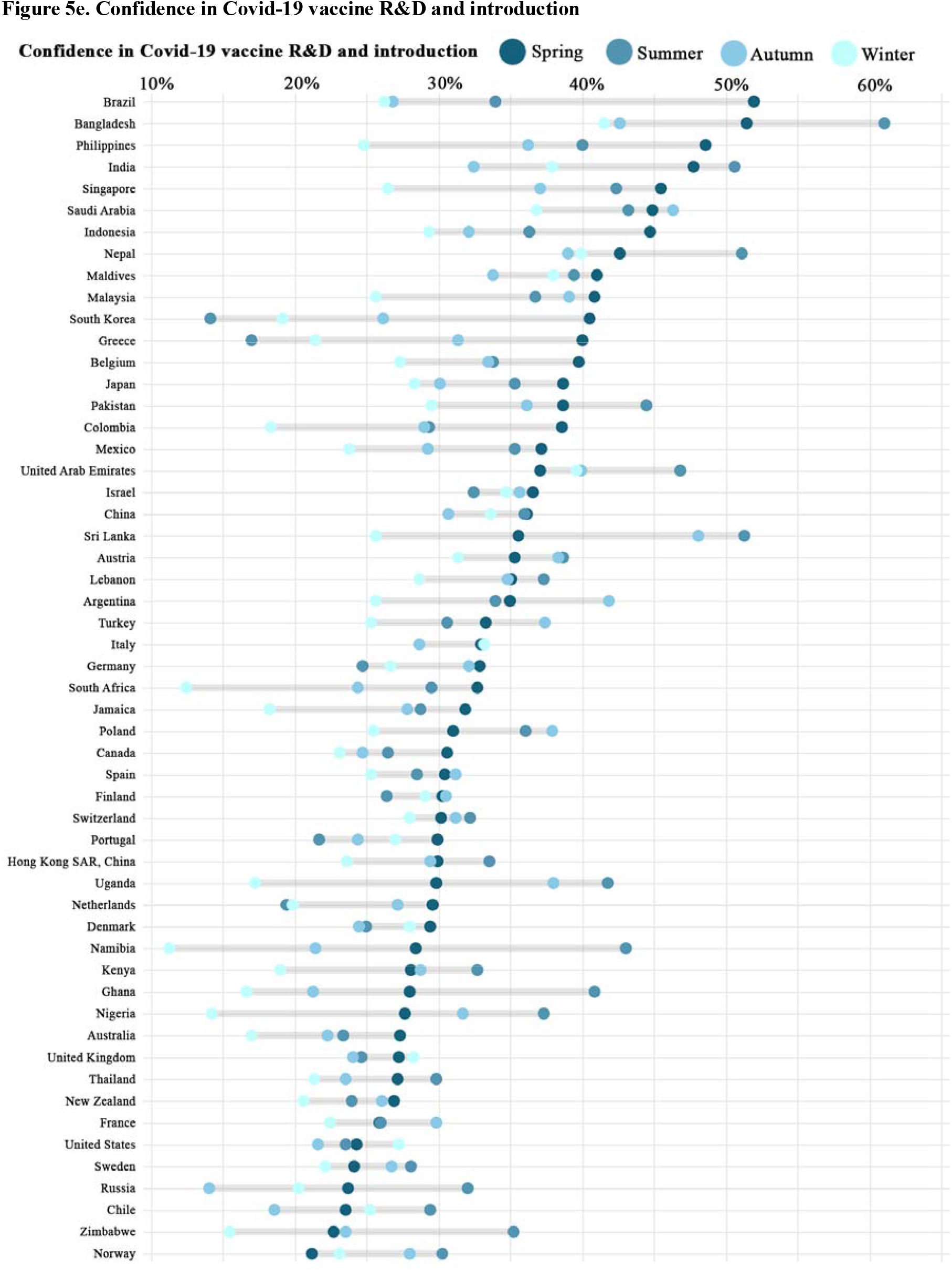
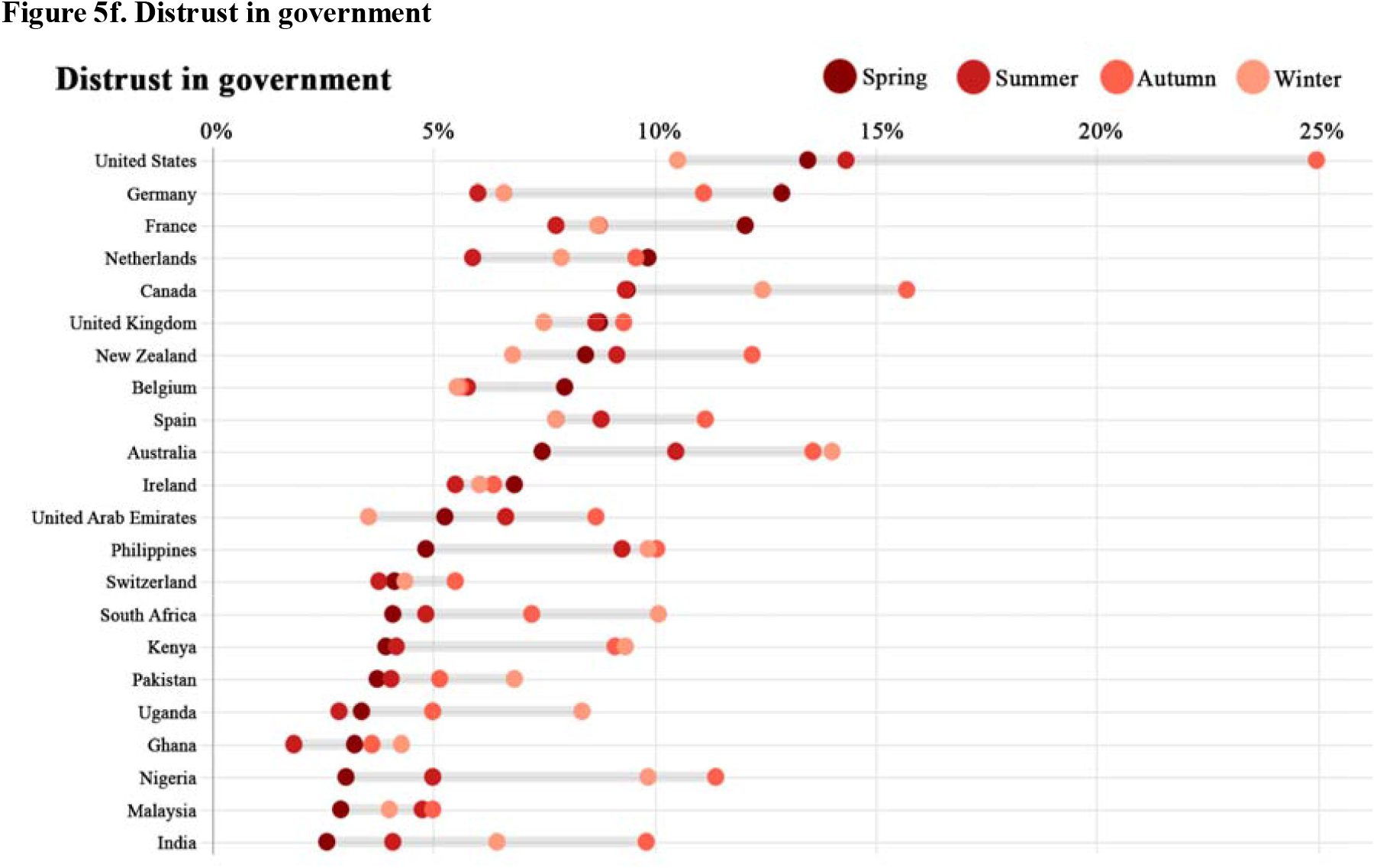
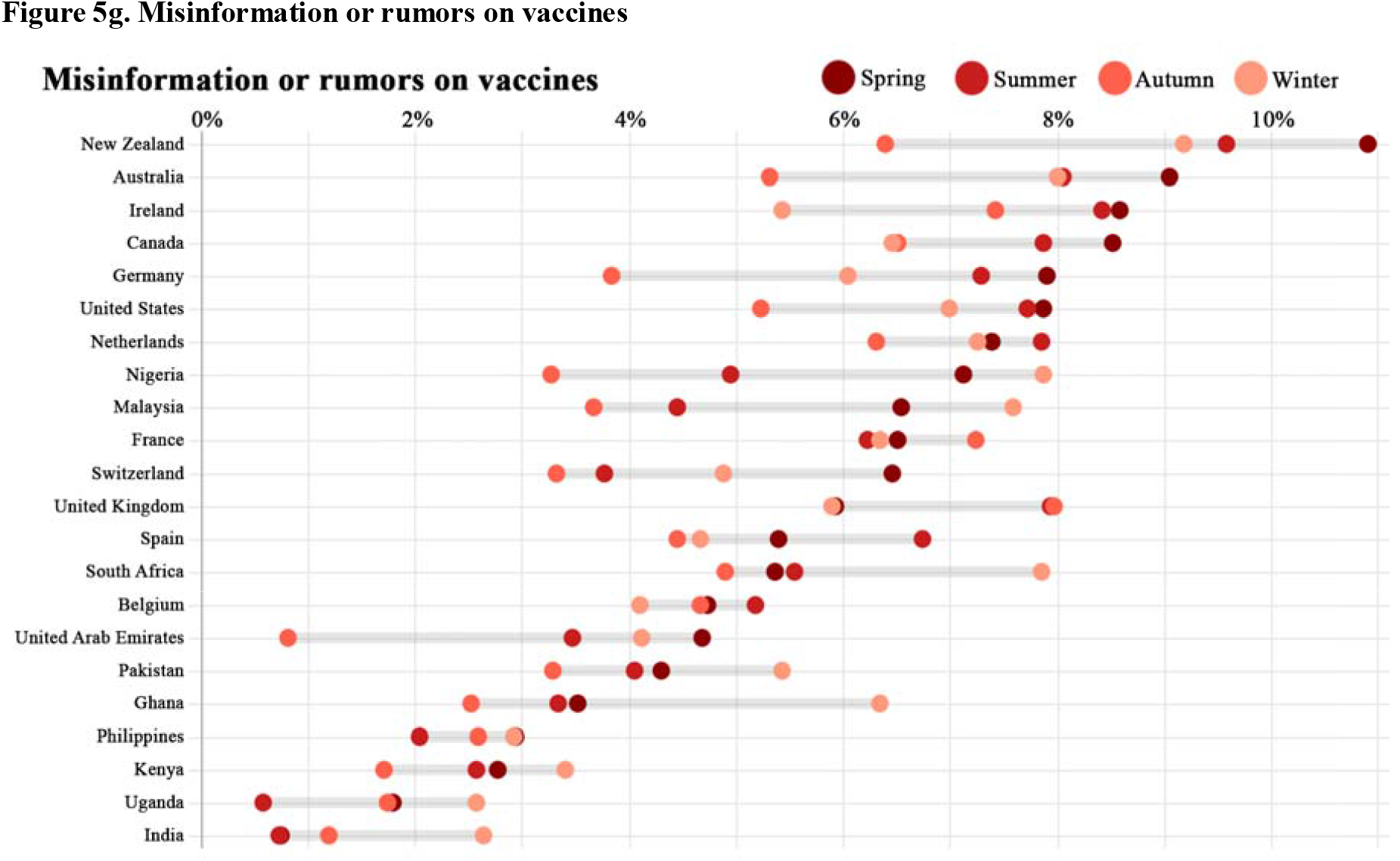
Country-level seasonal changes of Covid-19 vaccination intention and confidence between January 2020 and February 2021. At the country level, the graphs below illustrate the proportion of tweets which contain the following categories regarding Covid-19 vaccination intention or confidence during each season. Spring: March to May 2020; Summer: June to August 2020; Autumn: September to November 2020; Winter: December 2020 to February 2021. Countries are ranked based on each indicator’s values in the spring season.

Among all countries with sufficient data, the average proportion of tweets indicating Covid-19 vaccine being effective decreased from 11.99% in spring to 7.14% in winter, and the average proportion of tweets indicating Covid-19 vaccine being unsafe increased from 4.70% to 9.25% (Figure 5). For example, in Malaysia and Philippines, around 20% of tweets indicating Covid-19 vaccine being effective in spring, but the proportion decreased to around 7% in winter. In the Netherlands, proportion of tweets indicating Covid-19 vaccine being unsafe experienced a huge increase from 6.24% in spring to 13.85% in winter. In most countries, the prevalence of distrust in government increased, but the prevalence of misinformation or rumors decreased in tweets.

By country-level linear regressions, significant correlations were observed between intent to accept Covid-19 vaccination and vaccine confidence (Table 2 and appendix figure 5). We also validated our social media data with the previous survey data^1^, finding a positive linear correlation between intent to accept Covid-19 vaccination in our data and Covid-19 vaccine acceptance in the previous surveys (coefficient=0.16 in linear regression, p=0.064, R^2^=0.12; appendix figure 6).

**Table 2.** Country-level linear regressions between intent to accept Covid-19 vaccination and vaccine confidence or survey data.

| Measurements | Coefficient | 95% CI | p value | R<sup>2</sup> |
| --- | --- | --- | --- | --- |
| Internal measurements from Twitter data |  |  |  |  |
| Intent to reject Covid-19 vaccination | -3.96 | [-4.61, -3.32] | <0.001 | 0.74 |
| Belief that Covid-19 vaccine is effective | 2.17 | [1.36, 3.00] | <0.001 | 0.35 |
| Belief that Covid-19 vaccine is unsafe | -2.29 | [-2.91, -1.67] | <0.001 | 0.51 |
| Confidence in Covid-19 vaccine R&D and introduction | 1.08 | [0.95, 1.21] | <0.001 | 0.72 |
| Distrust in government | -1.72 | [-2.54, -0.89] | <0.001 | 0.25 |
| Misinformation or rumors on vaccines | -2.51 | [-3.24, -1.78] | <0.001 | 0.47 |
| Validation with survey data |  |  |  |  |
| Covid-19 vaccine acceptance survey* | 0.16 | [-0.01, 0.33] | 0.064 | 0.12 |

Co-efficient, slope (95% CI), p-value, and R<sup>2</sup> are from univariate linear regression. Dependent variable: proportion of tweets indicating intent to accept Covid-19 vaccination, for countries with relevant tweets >100.
\* Data from Wouters OJ, et al. Challenges in ensuring global access to COVID-19 vaccines: production, affordability, allocation, and deployment. The Lancet. 2021 Feb 12.

We further compared public engagement across different categories regarding Covid-19 vaccination intention and confidence (Table 3). On average, Covid-19 vaccine related tweets received 12.94 favorites, 2.55 retweets, 0.91 replies, and 0.28 quotes. Negative tweets, especially misinformation or rumors, received more engagement but had fewer followers than positive tweets. Even if they were sent by accounts that were less commonly followed, tweets containing misinformation or rumors received the highest number of favorites (17.98), the second highest replies (1.16) and quotes (0.37), and the third highest retweets (3.00); tweets indicating intent to reject Covid-19 vaccination received the highest replies (1.45) and quotes (0.41); tweets indicating distrust in government and belief that Covid-19 vaccine is unsafe were the most commonly retweeted (3.26 and 3.07 respectively). In contrast, tweets indicating intent to accept Covid-19 vaccination, belief that Covid-19 vaccine is effective, and confidence in Covid-19 vaccine R&D and introduction were less retweeted, replied to, and quoted.

**Table 3.** Public engagement in Covid-19 vaccine related tweets.

|  | **Follower** | **Retweet** | **Favorite** | **Reply** | **Quote** |
| --- | --- | --- | --- | --- | --- |
| All tweets | 2502.85(20737.55) | 2.55(194.20) | 12.94(1307.94) | 0.91(23.40) | 0.28(13.84) |
| Intent to accept Covid-19 vaccination | 2511.37(21878.67) | 2.44(186.15) | 15.24(1636.72) | 0.84(13.87) | 0.24(14.99) |
| Intent to reject Covid-19 vaccination | 2224.92(16224.20) | 2.26(82.22) | 8.40(310.48) | 1.45(81.98) | 0.41(15.03) |
| Belief that Covid-19 vaccine is effective | 2583.09(22342.67) | 1.66(87.84) | 11.03(708.83) | 0.81(9.59) | 0.18(12.44) |
| Belief that Covid-19 vaccine is unsafe | 2137.65(12929.20) | 3.07(210.00) | 11.99(1052.35) | 0.87(7.98) | 0.34(12.26) |
| Confidence in Covid-19 vaccine R&D and introduction | 2511.45(21297.92) | 1.97(154.43) | 14.09(968.68) | 0.80(11.11) | 0.22(9.11) |
| Distrust in government | 2891.09(20814.53) | 3.26(155.91) | 11.77(664.82) | 0.87(15.83) | 0.30(10.28) |
| Misinformation or rumors on vaccines | 2293.98(20954.31) | 3.00(173.09) | 17.98(1228.41) | 1.16(20.42) | 0.37(17.88) |

Mean (Standard Deviation) are shown.

## Discussion

To our knowledge, this is the largest global monitoring study of Covid-19 vaccination intention and confidence with social media data. We explore Covid-19 vaccination beliefs in over 100 countries from the beginning of the pandemic to February 2021, and refined deep learning-based classification helps achieve the temporal and spatial analyses of the evolving beliefs to Covid-19 vaccines across the world in real time. This global real-time surveillance study highlights the importance of regular social media monitoring to detect emerging trends to inform timely interventions to address Covid-19 vaccine hesitancy.

According to temporal analysis, acceptance and confidence towards Covid-19 vaccination have experienced a declining trend since the beginning of the pandemic in all world regions, with some regions recovering recently, though not to their original levels. The global declining trend in vaccine acceptance has been documented in previous surveys^4,20,21^, and a systematic review^3^ summarized these studies concluding that intention to accept Covid-19 vaccines decreased from >70% in March to <50% in October, 2020 around the world.

We detected a recent recovery of vaccine acceptance in several countries, most notably in the US and UK (as well as other countries in EUR), likely as a result of the reported high efficacy from vaccine trials and vaccine rollout in these countries. Consistent with our findings, household panel surveys in the US showed that vaccination intention increased from 39.4% in September to 49.1% in December 2020^22^. In the UK, 86% of adults who were reluctant to vaccinate against COVID-19 in December 2020 planned on accepting vaccines in February 2021^23^.

It should be noted that the global rise in Covid-19 vaccine acceptance since late 2020 was largely driven by the US and EUR. In contrast, other regions, especially AFR, experienced the declining trends in vaccine acceptance during 2020. Residents of low- and middle-income countries were more concerned with vaccine accessibility than their peers in high-income countries^19,24^. And we also found that many AFR countries worried more about vaccine effectiveness than others. Unequal access to Covid-19 vaccines and concerns on vaccine effectiveness may contribute to the low and declining acceptance and confidence in AFR^25^. To maintain vaccine acceptance worldwide, there is a need for strengthened international partnerships and coordination to ensure equitable access to Covid-19 vaccines globally^1^, and communication on vaccine effectiveness^25^.

Through spatial analysis, we found that intent to accept and confidence of Covid-19 vaccination were relatively high in SEAR, EMR and WPR, but low in AMR, EUR and AFR, in line with previous global surveys on Covid-19 and general vaccines^2,3,20,21^. For example, we found the highest acceptance in Bangladesh (76.00%) and India (72.29%) but low confidence across EUR in Covid-19 vaccine, which were usually found in global surveys on general vaccines^26^. Regional variation of Covid-19 vaccine acceptance also seemed similar to the use of facemasks which was prevalent in WPR countries such as Japan and China, but not in EUR and AMR^27^. The public in individual regions tend to percept similarly in Covid-19 vaccines and non-pharmaceutical interventions, influenced by societal and cultural paradigms^28^. However, vaccine hesitancy and premature relaxation of non-pharmaceutical interventions in individual countries may lead to the resurgence of Covid-19^29^. Covid-19 cases have resurged during March 2021 in the US and many EUR countries despite the introduction of Covid-19 vaccines, necessitating a new wave of lockdowns in some EUR countries.

In public engagement analysis, negative tweets, especially misinformation or rumors, were more engaged and discussed online by a small group than positive tweets. Information tends to become shorter, gradually inaccurate, and increasingly dissimilar between chains when propagating through the diffusion chains, which increases inaccuracy and misinformation on social media interaction^30^. Social media may be a double-edged sword that can disseminate health knowledge directly to the public, but also spread misinformation^10,31^. Their users suffered from online misinformation and exposure to anti-vaccine media, and anti-vaccine activities on social media have led to vaccine hesitancy and preventable deaths globally^24,32^. Policymakers and professionals should pay attention to online misinformation or rumors, and build more proactive public health responses on social media to promote dissemination and public engagement in accurate information.

Globally, we have shown a strong correlation between vaccine acceptance and beliefs about safety and efficacy, distrust in government, and misinformation. A recent study^33^ showed that, relative to factual information, misinformation led to a decline of 6 percentage points in Covid-19 vaccination intention in the UK and US, and scientific-sounding misinformation seemed more strongly associated with declines in vaccination intention. Our results were also consistent with the finding for general vaccines that the spread of misinformation or rumors on social media was significantly associated with vaccine hesitancy^34^. Therefore, campaigns about vaccine education should be strengthened to convince people of the safety and effectiveness of Covid-19 vaccines, and to address vaccine hesitancy in every country.

In addition, each region or country has its own features regarding Covid-19 vaccination intention and confidence. We found that many AFR countries worried more about vaccine effectiveness, while EUR, AMR, and WPR concerned more about vaccine safety (most concerns in Greece). Online misinformation or rumors were widespread in AMR, EUR, and South Korea (ranks first globally), and distrust in government was more prevalent in AMR (most prevalent distrust in the US). Therefore, targeted campaign strategies should be designed to improve vaccine acceptance in individual regions or countries.

### Strengths and limitations

The extensiveness, timeliness and accessibility of social media data, as well as the efficiency and accuracy of the pretrained deep learning models we used, provide a quick and effective method for detecting vaccination intention and confidence, especially in settings with limited sources and urgent timelines. This study could monitor populations who may not be represented in traditional surveys, and real-time social media data can eliminate reporting biases when speaking to a researcher^18^. Our findings can also be used as a reference point for survey data on a single country’s Covid-19 vaccination intention in the future.

This study has several limitations. First, although tweet data reflect the opinions of all Twitter users, such users may not be representative of the general population of the country. They are generally younger and more literate^35^. There may also be response biases since users may express their opinions differently online compared to in person. These biases are shared among all social media studies. Second, to ensure high performance of deep learning models, we did not train multilingual models. Our model only analyzed English tweets, reducing the representativeness for non-English speaking countries. Multilingual deep learning models are needed in other languages.

Despite these limitations, our data and methods provide a feasible approach to gaining valuable insight into an urgent area of research. Where traditional surveys are available, these produce similar results to ours, strengthening our confidence in the robustness of our study.

## Data Availability

Data described in this study can be found in appendix 2. The processed tweets dataset (~5.04 million, deidentified) is available on GitHub (https://github.com/xinyuuzhou/covid-19_vaccine_tweet_dataset). All code and deidentified raw data would be available on reasonable request (to replicate this study, or for further analysis, etc.) by contacting Zhiyuan Hou or Xinyu Zhou.

## Contributors

ZH and XZ contributed equally. ZH and XZ conceived and designed the study. XZ collected Twitter data, trained deep learning models, implemented the statistical analyses, and created the figures. ZH wrote the manuscript, and XZ and QX co-drafted some parts of the first version of the manuscript. All authors contributed to data interpretation, and finalised and approved the manuscript.

## Data sharing

Data described in this study can be found in appendix 2. The processed tweets dataset (∼5.04 million, deidentified) is available on GitHub (https://github.com/xinyuuzhou/covid-19_vaccine_tweet_dataset). All code and deidentified raw data would be available on reasonable request (to replicate this study, or for further analysis, etc.) by contacting Zhiyuan Hou or Xinyu Zhou.

## Acknowledgement

Zhiyuan Hou acknowledges financial support from the National Natural Science Foundation of China (No. 71874034), the National Key R&D Program of China (No. 2018YFC1312600 and 2018YFC1312604), and the National Institute for Health Research (EPIDZL9012) using UK aid from the UK Government to support global health research. We are thankful to Chen Wang from Software School of Fudan University for his helpful suggestions on fine-tuning deep learning models; Linyao Lu from School of Public Health, Fudan University for her help on literature review; and Yixing Tong and Fanxing Du from School of Public Health, Fudan University for their help on data coding analysis.

