## Appendix 1 for "Monitoring global trends in Covid-19 vaccination intention and confidence: a social media-based deep learning study"

**Details of the methodology**

**Details of rule-based removal of tweets**

We applied the following rules to exclude irrelevant tweets, such as tweets from news media, organizational accounts, or verified users. Note that after rule-based removal, there would be a much more thorough irrelevant tweet exclusion with the deep learning model.

- remove all tweets sent by verified users;
- remove tweets starts with “#COVID19 #vaccine news”, “#Breaking:”, “'#BREAKING:'”, “'BREAKING'”, “NEWS”, and “EXCLUSIVE”;
- remove tweet if we find “news” or “latest” in the user’s profile description;
- remove duplicated tweets;
- replace names of city, such as “New York”, with “the city”, and replace names of country, such as “United States”, with “the country”, to decrease country-level bias;
- remove tweets without content.

Additionally, note that before rule-based removal, we applied the strategy below to preprocess tweets:

- remove “\n”, links, and mentions (@username) inside tweets’ text.

**Details of the methodology**

**F_1_-score, hyperparameters, and performance of deep learning models**

F_1_-score is widely used to measure the performance of machine learning models. A high f_1_-score means the model not only reaches high positive predictive value (*accuracy or precision*), but also a high true positive rate (*recall*).

$$f_{1}-score= 2\cdot\frac{1}{\frac{1}{accuracy}+\frac{1}{recall}}= 2\cdot\frac{accuracy\cdot recall}{accuracy+recall}$$

Where,

$$accuracy= \frac{True Positive}{True Positive+False Positive}$$

$$recall= \frac{True Positive}{True Positive+False Negative}$$

Training seven categories in one model would limit the model’s performance. Therefore, in this study, we trained the following five deep learning models by fine-tuning CT-BERT ("digitalepidemiologylab/covid-twitter-bert-v2").

Model (1) is a binary classification model designed to classify tweets regarding “intent to accept Covid-19 vaccination”; Hyperparameters: max length=96, batch size=32, optimizer: AdamW, lr=1.5e-5, weight decay=1e-4, epoch=4;

Model (2) is another binary classification model to classify tweets regarding “intent to reject Covid-19 vaccination”; Hyperparameters: max length=96, batch size=32, optimizer: AdamW, lr=1.7e-5, weight decay=1e-4, epoch=3;

Model (3) is a multi-classifier for tweets about “belief that Covid-19 vaccine is effective”, “misinformation or rumors on vaccines”, “confidence in Covid-19 vaccine R&D and introduction”; Hyperparameters: max length=96, batch size=32, optimizer: AdamW, lr=3.1e-5, weight decay=5e-4, epoch=6;

Model (4) is another multi-classifier for tweets surrounding “belief that Covid-19 vaccine is unsafe”, and “distrust in government”; Hyperparameters: max length=96, batch size=32, optimizer: AdamW, lr=3.1e-5, weight decay=1e-4, epoch=5;

Model (5) is a binary classifier to remove irrelevant tweets, such as news, advertisements, and government announcements; Hyperparameters: max length=96, batch size=32, optimizer: AdamW, lr=1.7e-5, weight decay=1e-4, epoch=3.

After removing irrelevant tweets with model (5) (accuracy = 0.86, f_1_-score=0.89) and remove/preprocess tweets with the strategies above, we classified relevant tweets using model (1), (2), (3), and (4), and reached an overall accuracy at 0.82 and an overall f_1_-score at 0.81.

**Appendix Figure 1. Monthly trends of Covid-19 vaccination intention and confidence between January 2020 and February 2021 in selected countries.**

Monthly proportions of tweets expressing each category regarding Covid-19 vaccination intention or confidence and 95% Confidence Interval based on bootstrap sampling are shown from January 2020 to February 2021. Countries with sufficient tweets (N>2500) are shown.

Bootstrap sampling is a resampling method based on simple random sampling with replacement. In each resample, the standard setting is to randomly select N observations with replacement from the original N observations. Through taking a large number of (e.g. 1000) resamples, this technique can be used to estimate the sampling distribution of almost any statistic. It was implemented by Python library "Seaborn" using the default parameters.

**Appendix Figure 1a. Positive intention or confidence**

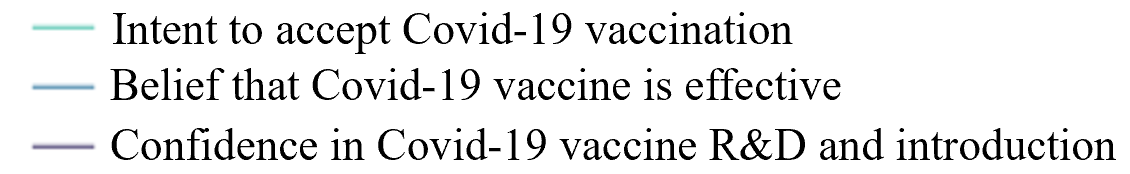

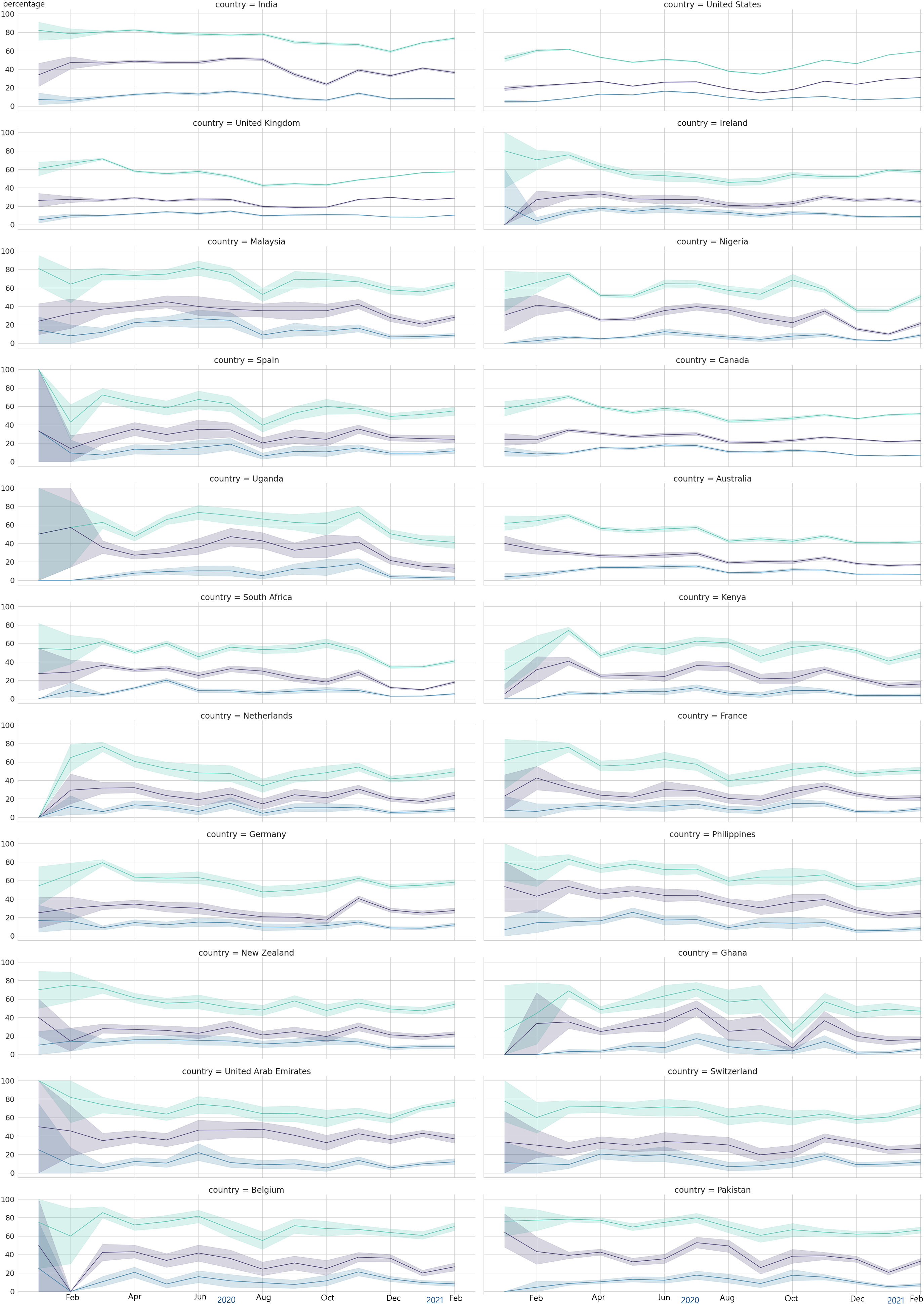

**Appendix Figure 1b. Negative intention or confidence**

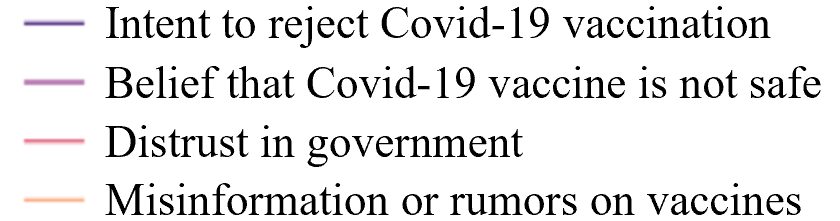

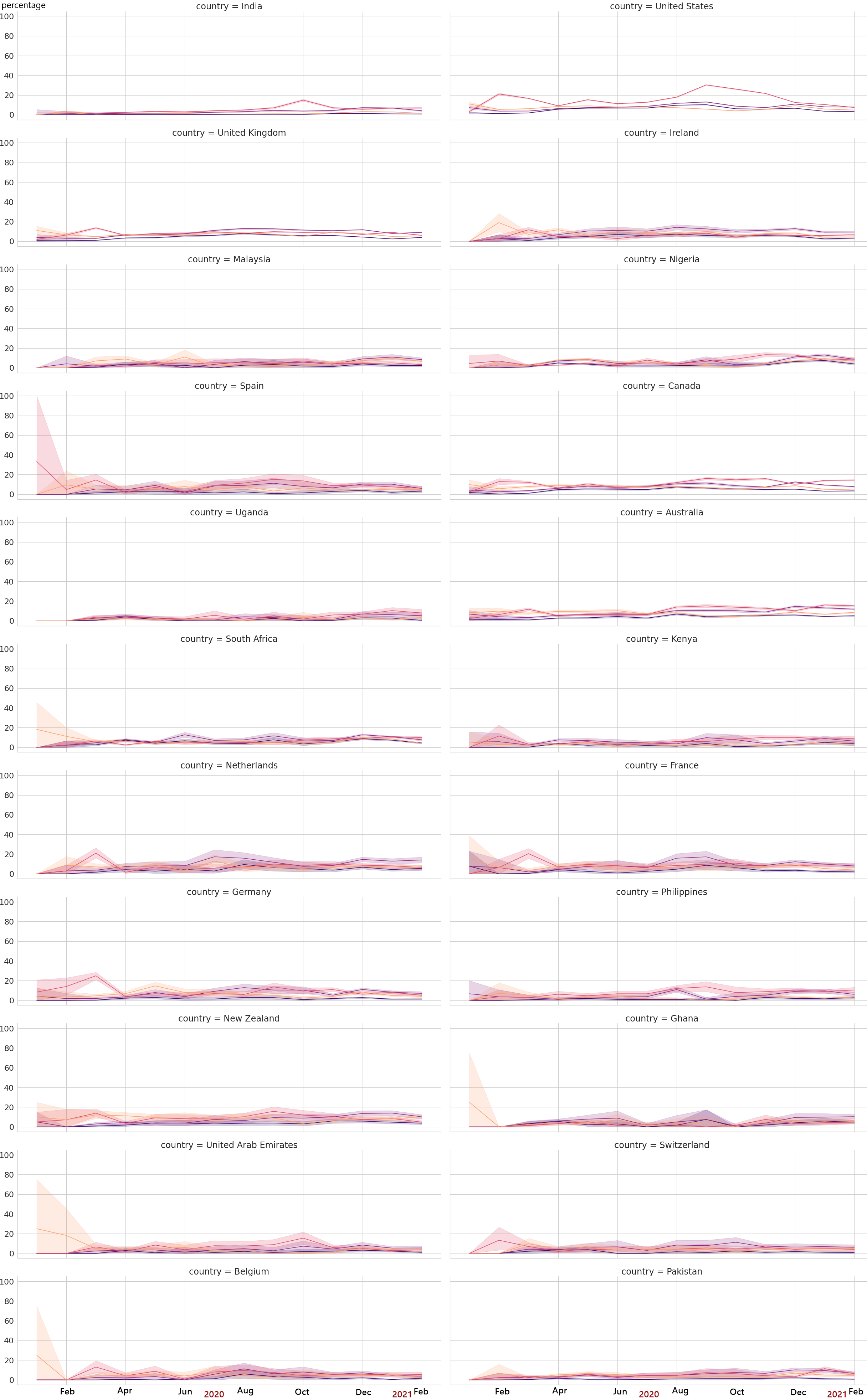

**Appendix Figure 2. Regional comparison of Covid-19 vaccination intention and confidence between January 2020 and February 2021.**

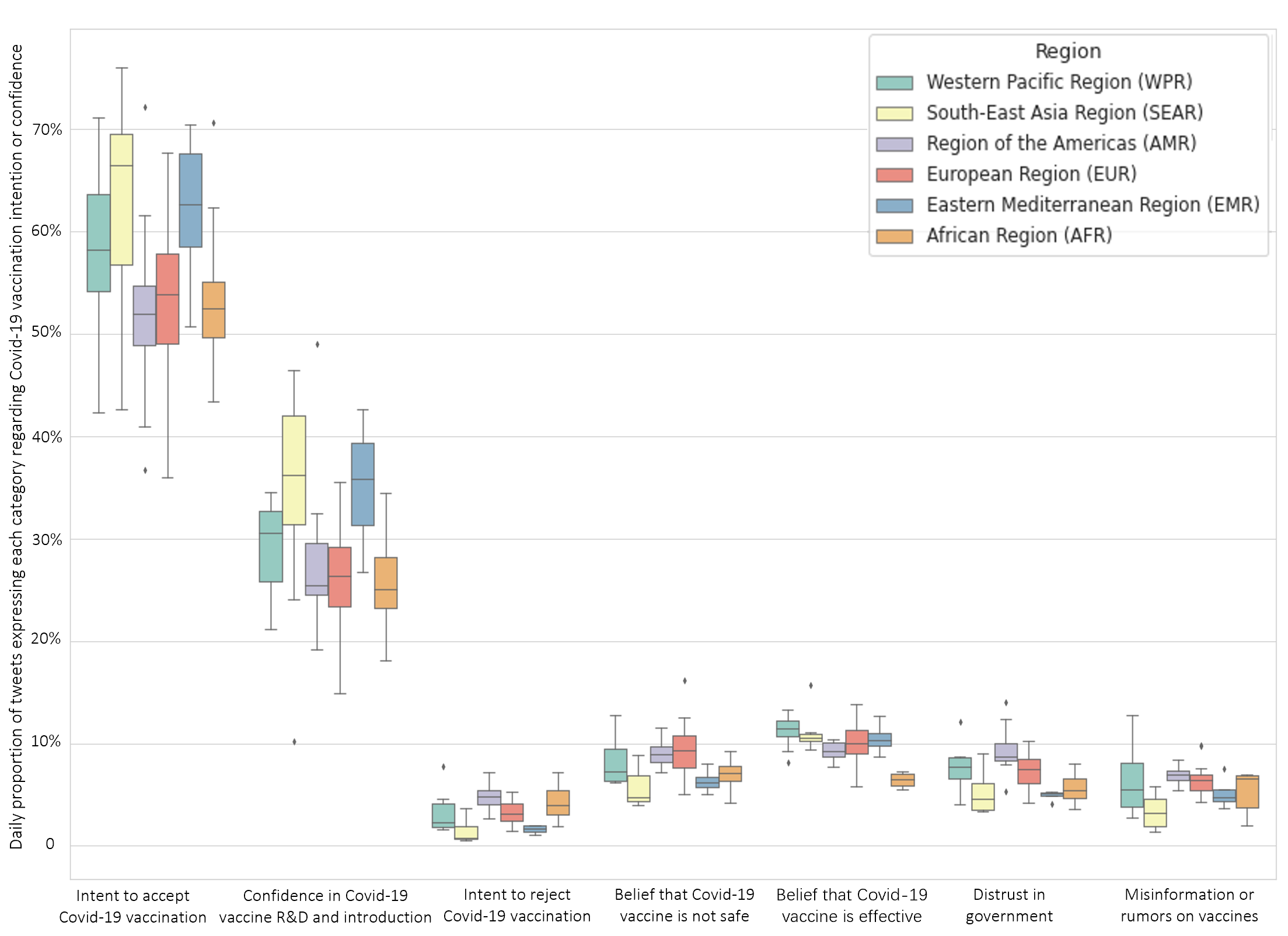

**Appendix Figure 3. Cross-country comparison of Covid-19 vaccination intention and confidence between January 2020 and February 2021.**

Proportions of tweets expressing each category regarding Covid-19 vaccination intention or confidence are shown.

**Appendix Figure 3a. Covid-19 vaccination intention**

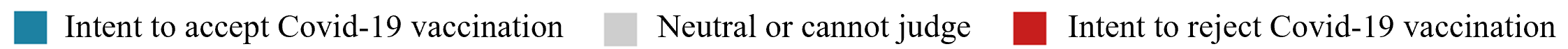

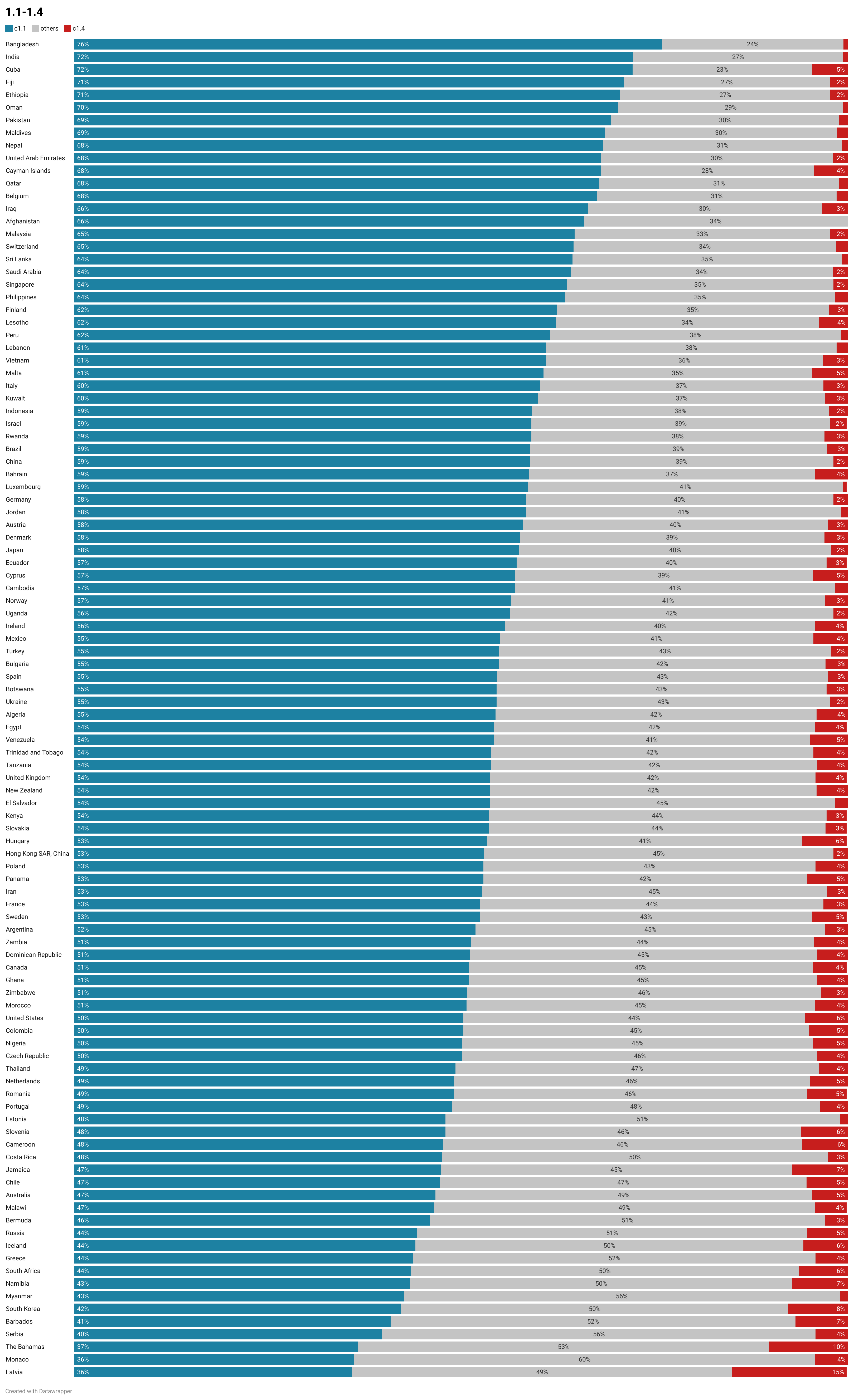

**Appendix Figure 3b. Covid-19 vaccination confidence**

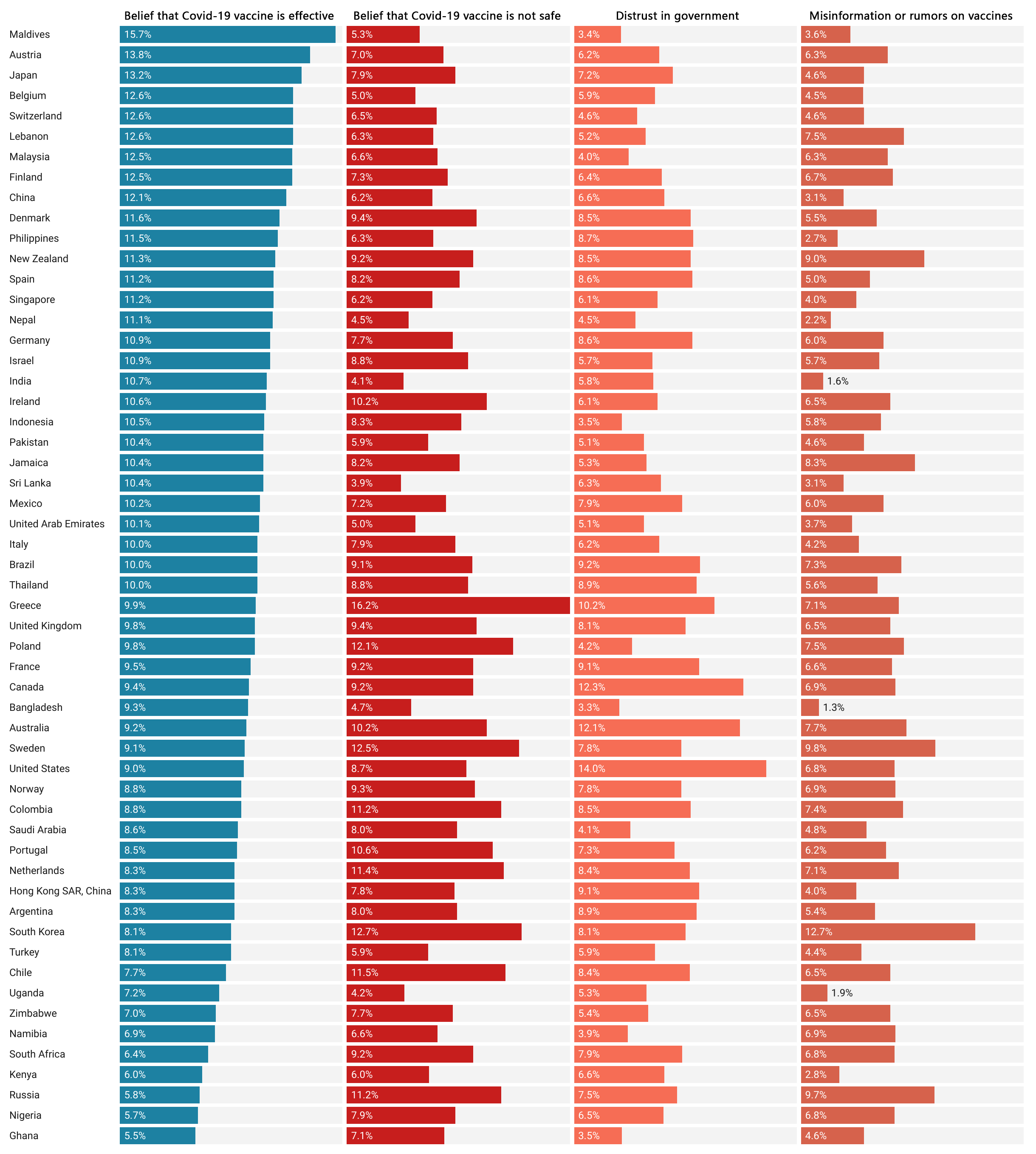

**Appendix Figure 3c. Confidence in Covid-19 vaccine R&D and introduction**

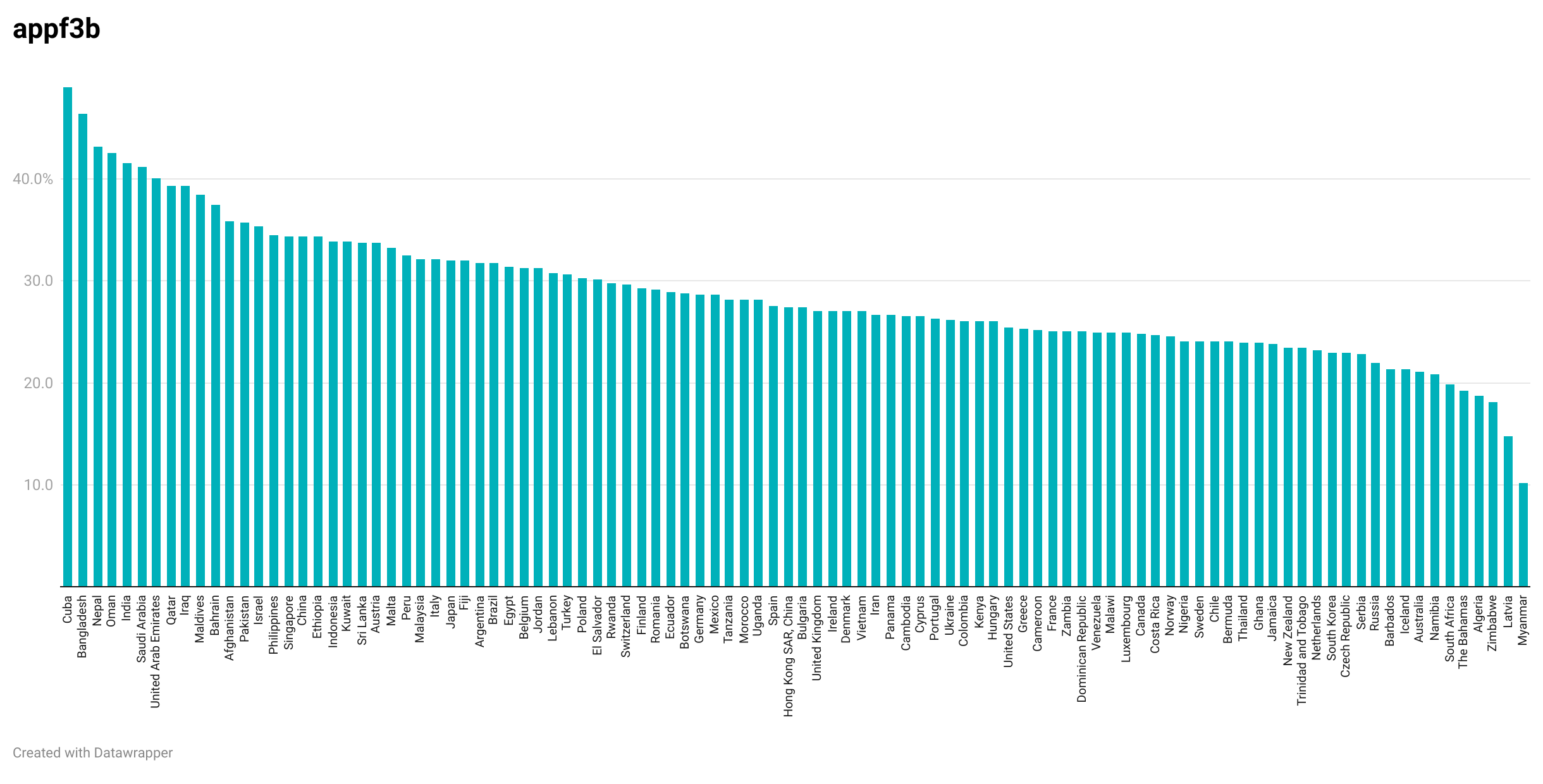

**Appendix Figure 4.** **Country-level monthly intention to accept Covid-19 vaccination between November 2020 and February 2021.**

Countries with sufficient tweets (N>1000) are shown.

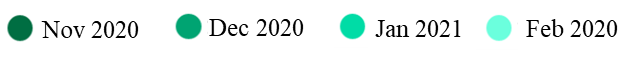

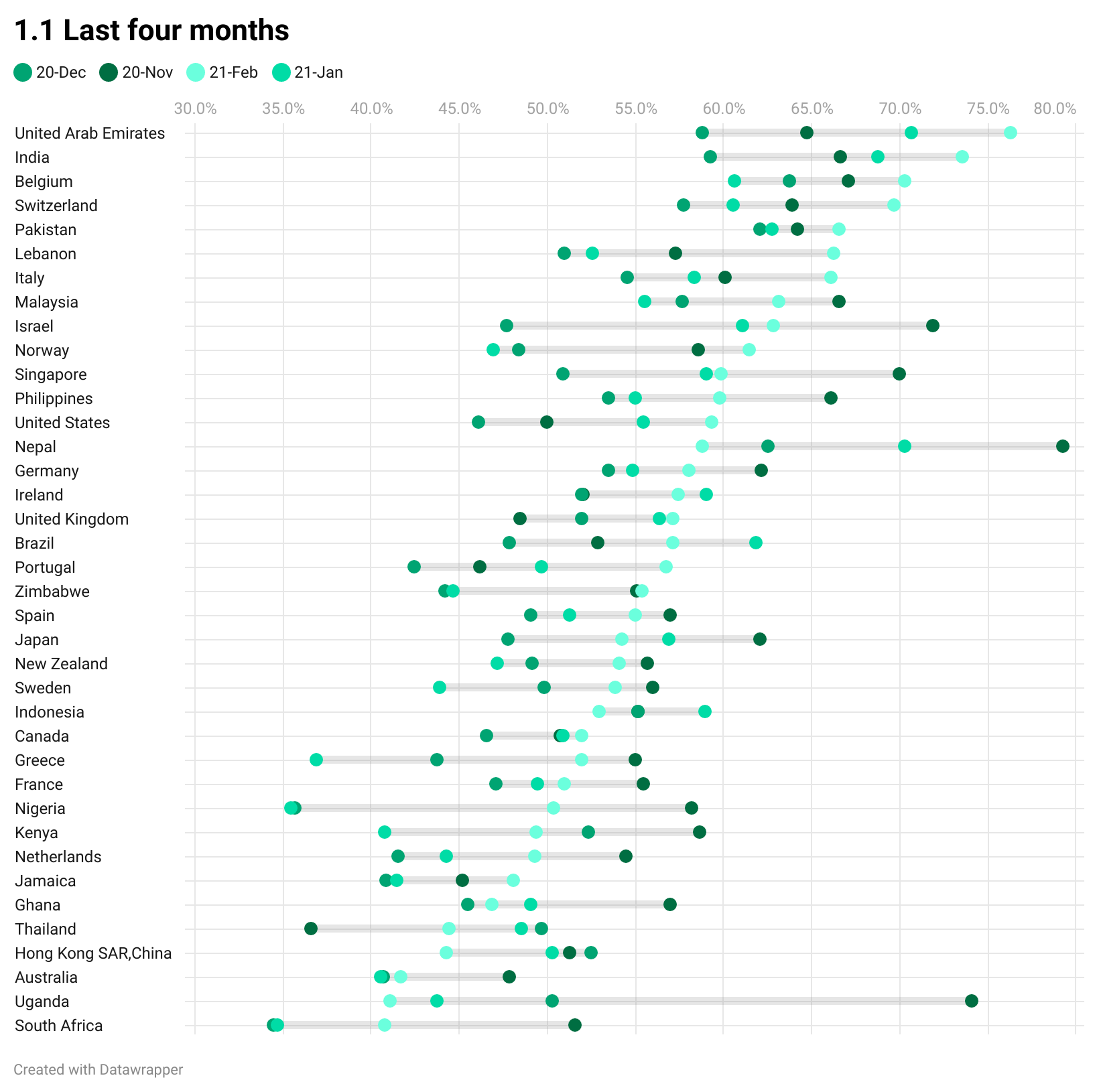

**Appendix Figure 5. Country-level correlation between intent to accept Covid-19 vaccination and intent to reject Covid-19 vaccine & Covid-19 vaccination confidence in Twitter.**

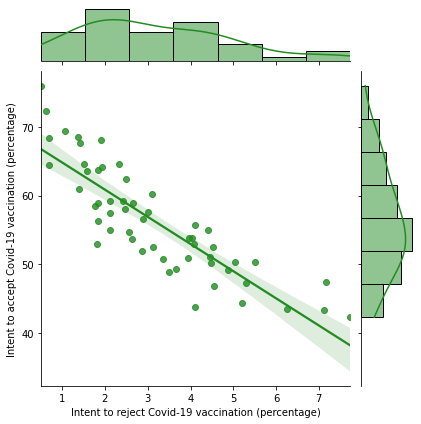

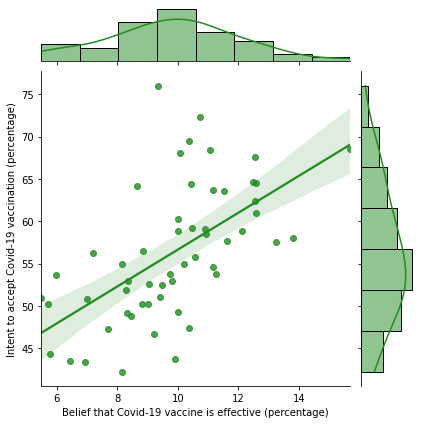

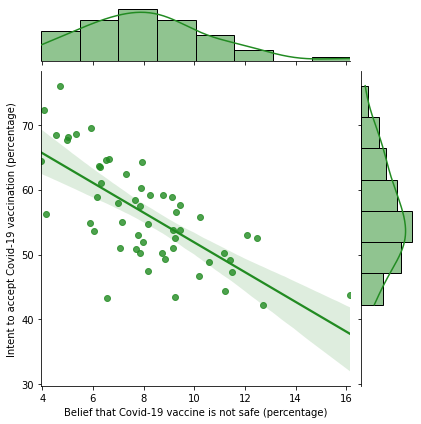

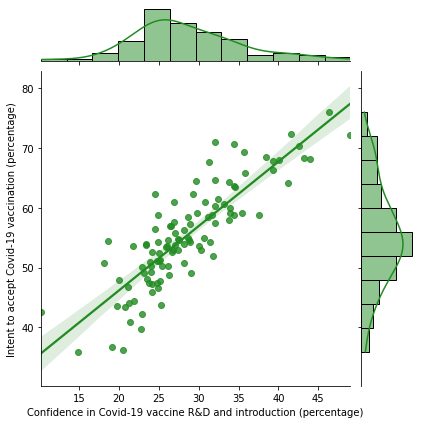

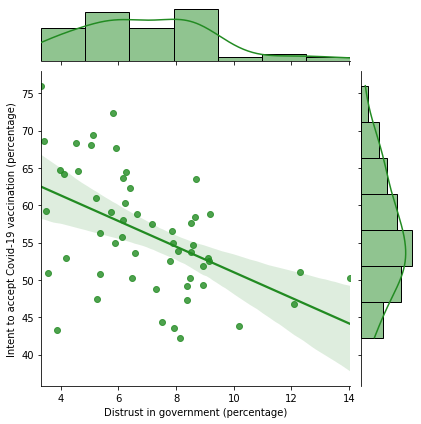

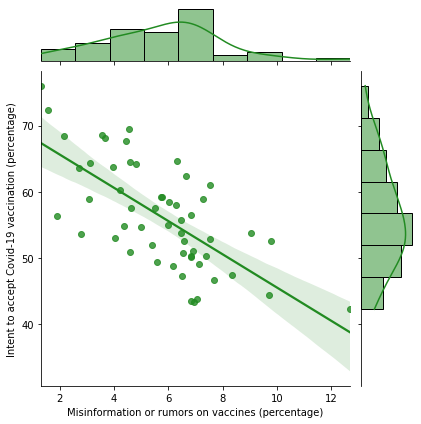

**Appendix Figure 6. Validation of social media data in our study with the previous global survey on Covid-19 vaccine acceptance.**

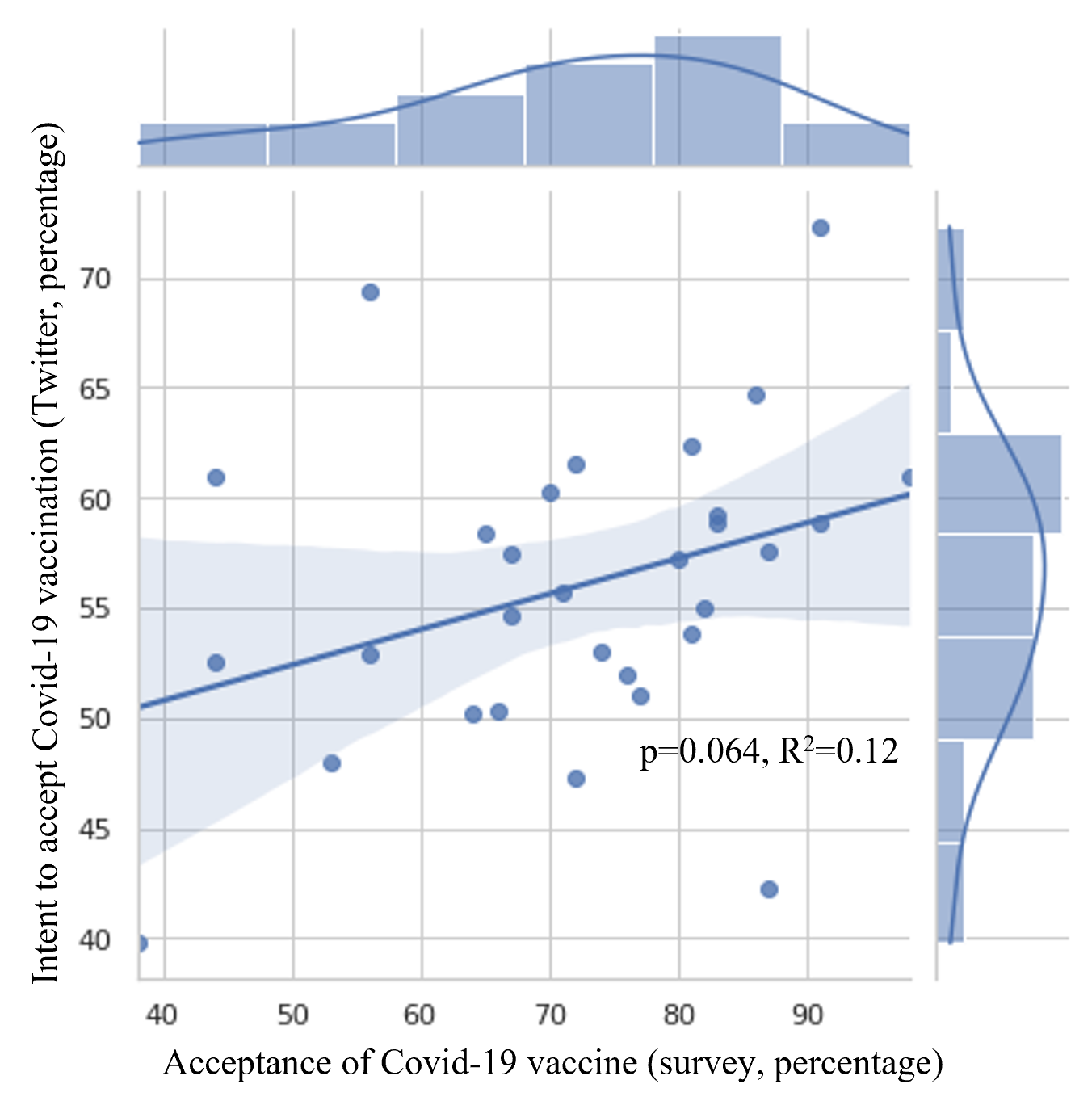

Survey data from Wouters OJ, et al. Challenges in ensuring global access to COVID-19 vaccines: production, affordability, allocation, and deployment. The Lancet. 2021 Feb 12.
